# Associations of insomnia on pregnancy and perinatal outcomes: Findings from Mendelian randomization and conventional observational studies in up to 356,069 women

**DOI:** 10.1101/2021.10.07.21264689

**Authors:** Qian Yang, M. Carolina Borges, Eleanor Sanderson, Maria C. Magnus, Fanny Kilpi, Paul J. Collings, Ana Luiza Soares, Jane West, Per Magnus, John Wright, Siri E. Håberg, Kate Tilling, Deborah A. Lawlor

**Affiliations:** MRC Integrative Epidemiology Unit at the University of Bristol, Bristol, UK; Population Health Sciences, Bristol Medical School, University of Bristol, UK; Centre for Fertility and Health, Norwegian Institute of Public Health, Oslo, Norway; Bradford Institute for Health Research, Bradford Teaching Hospitals NHS Foundation Trust, Bradford, UK; National Institute for Health Research Bristol Biomedical Centre, University Hospitals Bristol NHS Foundation Trust and University of Bristol, Bristol, UK

**Keywords:** insomnia, pregnancy and perinatal outcomes, Mendelian randomization, ALSPAC, MoBa

## Abstract

**Background:** Insomnia is common and associated with adverse pregnancy and perinatal outcomes in observational studies. Our aim was to test whether insomnia causes stillbirth, miscarriage, gestational diabetes, hypertensive disorders of pregnancy, perinatal depression, preterm birth, or low/high offspring birthweight (LBW/HBW).

**Methods and Findings:** We used two-sample Mendelian randomization (MR) with 81 single nucleotide polymorphisms instrumenting for a lifelong predisposition to insomnia. We used data (N=356,069) from the UK Biobank, FinnGen, and three European birth cohorts (Avon Longitudinal Study of Parents and Children (ALSPAC), Born in Bradford, and Norwegian Mother, Father and Child Cohort Study). Main MR analyses used inverse variance weighting (IVW), with weighted median and MR-Egger as sensitivity analyses. We compared MR estimates with multivariable regression of insomnia in pregnancy on outcomes in ALSPAC (N=11,745). IVW showed evidence of an effect of genetic susceptibility to insomnia on miscarriage (odds ratio (OR): 1.60, 95% confidence interval (CI): 1.18, 2.17), perinatal depression (OR 3.56, 95% CI: 1.49, 8.54) and LBW (OR 3.17, 95% CI: 1.69, 5.96). For other outcomes IVW indicated potentially clinically important adverse effects of insomnia (OR range 1.20 to 2.43), but CIs were wide and included the null. Weighted median and MR Egger results were directionally consistent, except for MR-Egger for gestational diabetes, perinatal depression, and preterm birth. Multivariable regression showed associations of insomnia at 18 weeks of gestation with miscarriage (OR 1.30, 95% CI: 1.12, 1.51), stillbirth (OR 2.10, 95% CI: 1.20, 3.69), and perinatal depression (OR 2.96, 95% CI: 2.42, 3.63), but not with LBW (OR 0.92, 95% CI: 0.69, 1.24). Key limitations are potential horizontal pleiotropy and low statistical power in MR, and residual confounding in multivariable regression.

**Conclusions:** There is evidence of causal effects of insomnia on miscarriage, perinatal depression, and LBW. We highlight the need for larger studies with genomic data and pregnancy outcomes.

**Author summary:** *Why was this study done?:*

- Insomnia in pregnancy was associated with higher risks of adverse pregnancy and perinatal outcomes in observational studies.
- It is currently no clear whether insomnia causes adverse pregnancy and perinatal outcomes or whether the unfavourable associations are explained by confounding.
- No Mendelian randomization has been conducted to explore the association of insomnia with adverse pregnancy and perinatal outcomes.

*What did the researchers do and find?:*

- We used data on up to 356,069 women from UK Biobank, FinnGen and three birth cohorts, and assessed whether genetic susceptibility to insomnia was associated with stillbirth, miscarriage, gestational diabetes, hypertensive disorders of pregnancy, perinatal depression, preterm birth, low offspring birthweight, and high offspring birthweight in two-sample Mendelian randomization.
- To triangulate with our Mendelian randomization estimates, we conducted multivariable regression in 11,745 women from the Avon Longitudinal Study of Parents and Children, where insomnia was measured in pregnancy.
- We found consistent evidence from Mendelian randomization and multivariable regression that insomnia was associated with higher risks miscarriage and perinatal depression, and Mendelian randomization also suggested an unfavourable effect on low offspring birthweight.

*What do these findings mean?:*

- Interventions to improve healthy sleep in women of reproductive age might be beneficial to a healthy pregnancy.

## Introduction

Insomnia, which affects approximately 10 to 20% of the adult population, is usually defined as a difficulty in getting to sleep or remaining asleep, or having a nonrestorative sleep, and such sleep impairment can be associated with daytime sleepiness [1, 2]. Physical and hormonal changes during pregnancy increase susceptibility to insomnia [3, 4].

Most evidence on the relationship between insomnia during pregnancy and adverse pregnancy and perinatal outcomes has come from observational studies. The most recently updated systematic reviews of observational studies suggest that pregnancy-related insomnia and poor sleep quality are associated with higher risks of gestational diabetes (GD) [5, 6], hypertensive disorders of pregnancy (HDP) [6], perinatal depression [7], and preterm birth (PTB) [6]. Other observational studies have shown that specific conditions that relate to insomnia are also associated with adverse pregnancy and perinatal outcomes. For example, meta-analyses combining four case-control studies showed that going to sleep in a supine position (plausibly exacerbating sleep-disordered breathing that contributes to decreased sleep quality [4, 8]) was associated with higher risk of stillbirth [9] and small-for-gestational age (SGA) [10]. Sleep-disordered breathing, obstructive sleep apnoea and restless legs syndrome have also been shown to associate with higher risks of GD, HDP, large-for-gestational age (LGA) and low offspring birthweight (LBW) [6]. However, it remains unclear whether insomnia causes adverse pregnancy outcomes or whether these associations are explained by confounding, e.g. due to socio-economic status and lifestyle factors. It is also possible that some of these studies reflect reverse causation. For example, all four studies included in the systematic review for perinatal depression were cross-sectional [7]. As disturbed sleep is a symptom of depression it is unclear whether these studies reflect a causal effect of insomnia, or it is part of the diagnostic criteria. Furthermore, most individual studies focus on just one or two outcomes. Examining potential effects on a range of adverse pregnancy and perinatal outcomes is important to understand the overall health impact of insomnia during pregnancy.

Three randomized control trials (RCTs) assessing the effects of interventions to prevent insomnia on adverse pregnancy and perinatal outcomes have been published [11-13]. All three of these used cognitive behavioural interventions targeted at reducing insomnia, with the primary outcome being Edinburgh Postnatal Depression Scale scores. The first RCT reported a difference in mean score of - 0.21 (95% confidence interval (CI): -0.30, -0.11) in 208 randomized women, with equivalent results for the other two being -0.24 (95% CI: -0.67, 0.19, N=194) and 0.34 (95% CI: -0.26, 0.93, N=91) [11-13]. The small number of RCTs, their small sample sizes, and directional inconsistency but overlapping CIs make it difficult to draw conclusions, and none of them explored other adverse pregnancy or perinatal outcomes.

Mendelian randomization (MR) provides an alternative way to assess the impact of insomnia on adverse pregnancy and perinatal outcomes by using genetic variants (mostly single nucleotide polymorphisms [SNPs]) as instrumental variables (IVs) for insomnia [14, 15]. MR is less prone to confounding than observational studies, as genetic variants are randomly allocated at meiosis and cannot be influenced by the wide range of socio-demographic or behavioural factors which conventionally confound observational studies, nor can they be influenced by health status [14, 15]. Under key assumptions (discussed in methods), MR can be used to estimate a causal effect from the associations of the SNPs with the exposure and outcome. In two-sample MR, the SNP-exposure and SNP-outcome associations are estimated using different studies [16]. This approach has previously been used to evaluate causal effects of self-reported insomnia on risks of type 2 diabetes [17, 18], hypertension [19] and cardiovascular disease [18, 20, 21] in non-pregnant populations, but not pregnancy and perinatal outcomes.

The aims of this study are to (I) explore the causal effects of maternal genetic susceptibility to insomnia on stillbirth, miscarriage, GD, HDP, perinatal depression, PTB, LBW, and high offspring birthweight (HBW), using two-sample MR, and (II) compare those findings with conventional multivariable regression analyses of self-reported insomnia during pregnancy with the same outcomes.

## Methods

### Study populations

This study was undertaken using data from the MR-PREG consortium, which aims to explore causes and consequences of different pregnancy and perinatal outcomes [22]. We used individual-level data from UKB women (N=208,140, recruited between 2006-2010), and mother-offspring pairs from ALSPAC (N=6,826, recruited between 1991-1992), BiB (N=2,940, recruited between 2007-2010) and MoBa (N=14,584, recruited between 1999-2009). To be comparable across all cohorts, only genetically unrelated women of European descent with qualified genotype data (and with singleton offspring in birth cohorts) were eligible for inclusion in our analyses (S1 Fig in S1 File). We also used summary-level genetic association data from FinnGen – the national wide network of Finnish biobanks (N=up to 123,579 women) [23]. All studies had ethical approval from relevant national or local bodies and participants provided written informed consent. Details of the recruitment, information on genetic data and measurements of baseline characteristics of each cohort are described in Supplementary Text (S1 File).

### Outcomes measures

We explored potential effects of insomnia on eight binary outcomes: ever experiencing stillbirth, ever experiencing miscarriage, GD, HDP, perinatal depression, PTB (gestational age <37 completed weeks), LBW (<2,500 grams) and HBW (>4,500 grams). Full details about how these outcomes were measured and derived in each participating study, and how we harmonised them across studies can be found in S1 Table (S2 File). We were not able to measure pre-eclampsia and gestational hypertension separately, because of the small number of definite cases of pre-eclampsia, and because of differences between studies in data collection and definitions.

In UKB gestational age was only available for a small subset of women (N=7280) who delivered a child during or after 1989, the earliest date for which linked hospital labour and perinatal data are available [24]. As a result, numbers with data on PTB are smaller than for any other outcome, and we *a priori* decided to examine associations with LBW and HBW rather than SGA and LGA. For most outcomes in UKB, women reported their experience retrospectively in a questionnaire completed at recruitment when they were aged 40-60 years.

In the three birth cohorts most outcomes were prospectively obtained (from self-report or clinical records) during an index pregnancy and the perinatal period. The two exceptions were history of stillbirth and miscarriage, which were retrospectively reported at the time of the index pregnancy when women were asked if they had ever experienced a (previous) stillbirth or miscarriage. We explored the possibility of examining associations with miscarriage and stillbirth in the index pregnancy. However, numbers were too small for reliable results in either MR or multivariable regression, and for miscarriage we were concerned about misclassification or selection bias due to women who had experienced a miscarriage prior to recruitment. If multiple pregnancies were enrolled in the birth cohorts, we randomly selected one pregnancy per woman.

Data from FinnGen were available for four of our outcomes: ever experiencing miscarriage, GD, HDP and PTB, which were defined based on International Classification of Diseases codes.

### Insomnia measures

Self-reported information on insomnia was obtained from two of the studies. In UKB information on lifetime insomnia was used to generate SNP-insomnia associations in women for use in MR analyses in UKB and the birth cohorts. ALSPAC collected data on insomnia during pregnancy, and this was used for conventional confounder-adjusted multivariable regression.

In UKB, insomnia was self-reported at recruitment via the question “Do you have trouble falling asleep at night or do you wake up in the middle of the night?” with responses “never/rarely”, “sometimes”, “usually” and “prefer not to answer”. For our analyses we collapsed these categories to generate a binary variable of usually experiencing insomnia (i.e. “usually” [cases] versus “sometimes” + “never/rarely” [controls]) as this was how the responses were categorised in the published genome-wide association study (GWAS) that we have used to select genetic IVs [18].

In ALSPAC, insomnia in pregnancy was self-reported, at 18 and 32 weeks of gestation, using the question “Can you get off to sleep alright?” with options “Very often”, “Often”, “Not very often” and “Never”. At each time point, we compared “Not very often” + “Never” [cases] versus “Very often” + “Often” [controls]. We acknowledge that the two studies are using different questions and that definitions of insomnia vary across published literature [2]. For ease of reading throughout the paper we refer to results reflecting genetic susceptibility to insomnia (MR) and reporting insomnia in pregnancy (multivariable regression).

### SNP selection and SNP-insomnia associations

To identify genetic IVs for insomnia, we searched the GWAS published between January 2017 and February 2021 on PubMed and Neale Lab website [25]. We found seven insomnia GWAS reporting genome-wide significant SNPs (details in S2 Table in S2 File). Of these we selected SNPs from the largest GWAS (total N = 709,986 women, 29% from UKB and 71% from 23andMe), which provided female-specific results [18]. This GWAS identified 83 loci containing 87 lead SNPs that were robustly associated with insomnia (P-value <5×10^−8^) after pooling UKB and 23andMe women together. We removed 6 SNPs that were correlated to other SNPs (*linkage disequilibrium*) at an R^2^ threshold of 0.01 or higher, based on all European samples from the 1000 genome project [26]. Associations (reported in log odds ratios [ORs]) of the remaining 81 lead SNPs from the women only GWAS were extracted and listed in S3 Table (S2 File).

We fitted linear regression to individual-level data from 208,140 UKB women to recalculate SNP-insomnia associations for two-sample MR analyses, to avoid non-collapsibility of ORs and difficult interpretation of MR estimates’ unit [27]. We adjusted the linear models for genotyping batch, top 40 principal components (PCs) and women’s age.

### SNP-outcome associations

We estimated the associations between maternal SNPs and outcomes (log odds ratio (OR) and standard errors) for each of the 81 insomnia-related SNPs. In UKB, we randomly separating women in half (giving two datasets, A and B) for our split cross-over two-sample MR [28], given UKB was also included in the GWAS of insomnia. We then estimated SNP-outcome associations in each split sample using logistic regression, adjusting for genotyping batch, top 40 PCs, and women’s age. In the birth cohorts, we estimated the SNP-outcome associations using logistic regression, adjusting for (I) top 20 PCs and women’s age in ALSPAC; (II) top 10 PCs and women’s age in BiB; and (III) genotyping batch, top 10 PCs and women’s age in MoBa. We extracted associations of the 81 SNPs with miscarriage (O15_ABORT_SPONTAN), GD (GEST_DIABETES), HDP (O15_GESTAT_HYPERT), and PTB (O15_PRETERM) from FinnGen, which were generated using SAIGE (mixed-effects logistic regression [29]) adjusting for genotyping batch, top 10 PCs and women’s age [23]. Then we meta-analysed those associations from ALSPAC, BiB, MoBa and FinnGen using fixed-effects with inverse variance weights. Three SNPs (i.e. rs10947428, rs9943753, and rs117037340) were excluded from BiB analyses due to their minor allele frequency lower than 1%.

### Assessment of confounders in ALSPAC for multivariable regression

We considered maternal age at time of delivery, education, body mass index at 12 weeks of gestation, smoking status in pregnancy, alcohol intake in the first three months of pregnancy and household occupational social class as potential confounders based on their known or plausible associations with maternal insomnia and pregnancy and perinatal outcomes. Details of confounders were based on maternal self-report, and are fully described in Supplementary Text (S1 File).

### Statistical analyses

#### Two-sample MR

As shown in Fig 1, we conducted two-sample MR analyses of maternal insomnia on pregnancy and perinatal outcomes. In UKB, we conducted a split cross-over two-sample MR [28]. Specifically, we used SNP-insomnia associations from dataset A and SNP-outcomes associations from dataset B (A on B) and vice-versa (B on A), and then meta-analysed the MR estimates from the two together for each insomnia-outcome pair using fixed-effects (with inverse variance weights). For the two-sample MR using the rest of the cohorts, we used SNP-insomnia associations from UKB women and the pooled SNP-outcome associations combining ALSPAC, BiB, MoBa and FinnGen. For each outcome, we pooled MR estimates from all cohorts using fixed-effects (with inverse variance weights), and used leave-one (study)-out analysis to assess the degree of heterogeneity between cohorts.

**Fig 1.**
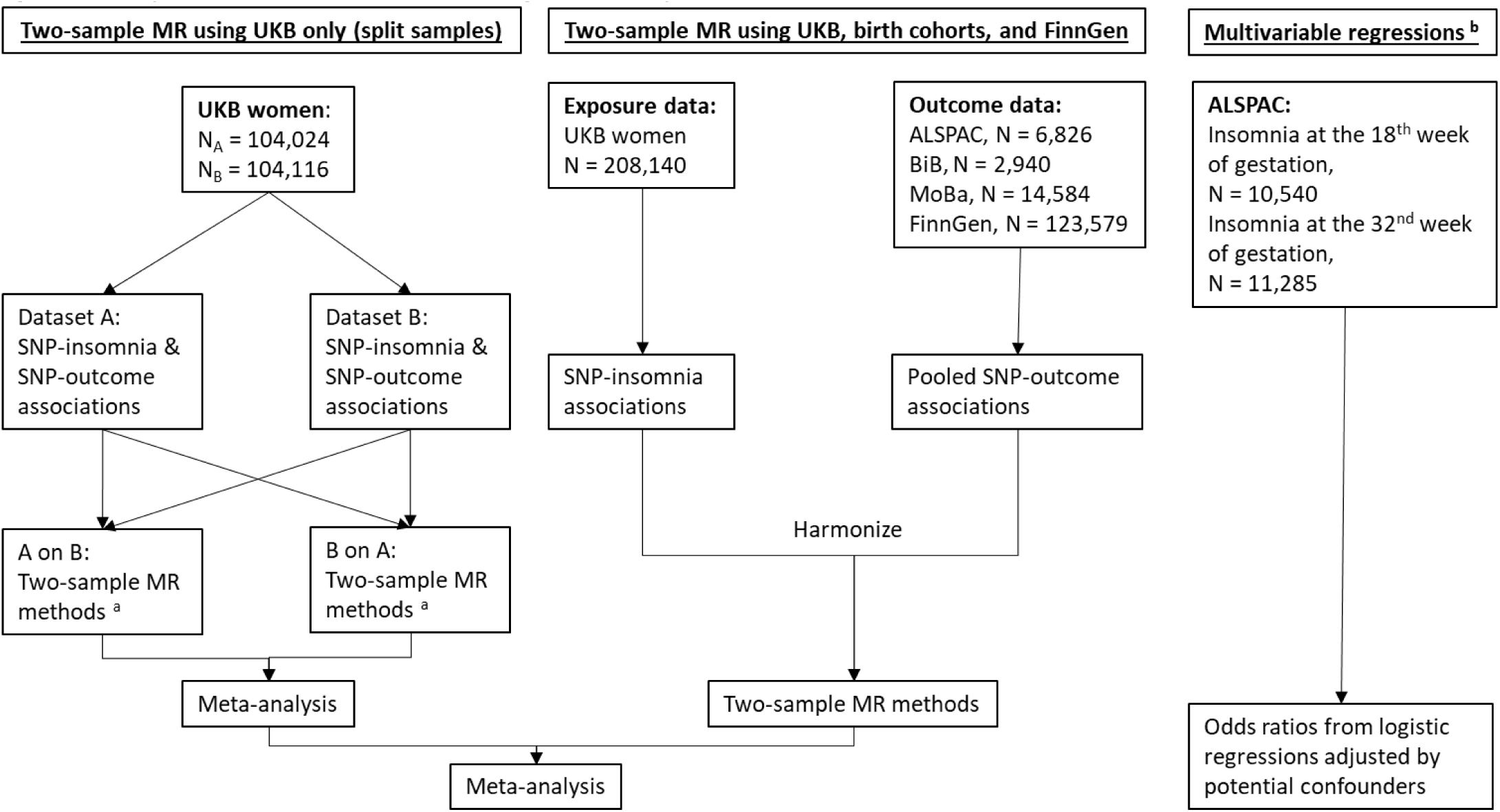
Summary of methods and data contributing to this study. ^a^ Two-sample MR methods include: IVW, MR-Egger, weighted median, and leave-one-out analysis. ^b^ Multivariable regression analysis adjusted for maternal age at time of delivery, social class, education, body mass index at 12 weeks of gestation, smoking status in pregnancy and alcohol intake in the first three months of ALSPAC pregnancy. Abbreviations: ALSPAC, Avon Longitudinal Study of Parents and Children; BiB, Born in Bradford; GWAS, genome-wide association study; IVW, inverse variance weighted; MoBa, Norwegian Mother, Father and Child Cohort Study; MR, Mendelian randomization; SNP, single nucleotide polymorphism; UKB, UK Biobank.

In the main analyses, we used the MR IVW method, which is a regression of the estimates for SNP-outcomes associations on SNP-insomnia associations weighted by the inverse of the SNP-outcome associations variances, with the intercept of the regression line forced through zero [30]. The IVW estimates should provide an unbiased estimate of a causal effect in the absence of unbalanced horizontal pleiotropy [30]. To explore potential unbalanced horizontal pleiotropy, our sensitivity analyses included (I) estimating between-SNP heterogeneity (which if present may be due to one or more SNPs having horizontal pleiotropic effects on the outcome) using Cochran’s Q-statistic and leave-one (SNP)-out analysis, and (II) undertaking analyses with weighted median [31] and MR-Egger [32], which are more likely to be robust in the presence of invalid IVs. The weighted median method is unbiased so long as less than 50% of the weight is from invalid instruments (i.e. if one SNP contributing more than 50% of the weight across the SNP-insomnia associations or several SNPs that contribute more than 50% introduce horizontal pleiotropy the effect estimate is likely to be biased) [31]. MR-Egger is similar to IVW except it does not constrain the regression line to go through zero; if the MR-Egger intercept is not null it suggests the presence of horizontal pleiotropy, and the MR-Egger slope provides an effect estimate corrected for unbalanced horizontal pleiotropy [32].

However, MR-Egger has considerably less statistical power than IVW. Further details of these MR methods are provided in our previous study [33]. When using MR to assess the effect of maternal exposures in pregnancy on offspring outcomes, results might be biased via a path from maternal genotypes to maternal/offspring outcomes due to fetal genotype [34]. To explore this, we compared SNP-outcome associations with versus without adjustments for fetal genotypes in the pooled birth cohort analyses.

We evaluated the strength of IVs using both proportion of variances of maternal insomnia explained by the 81 SNPs (R^2^) and F-statistic [35]. We selected SNPs robustly related to insomnia in the general female population rather than in pregnant women. Therefore, we explored associations of the 81 SNPs with woman’s insomnia measured at 18 and 32 weeks of gestation in ALSPAC using logistic regressions to determine whether those SNPs related similarly to insomnia in pregnancy. We adjusted for the top 20 PCs and women’s age.

#### Multivariable regressions in ALSPAC

In ALSPAC, we explored the observational associations of insomnia at 18 and 32 weeks of gestation with binary outcomes using logistic regression, with adjustment for measured confounders. We had to regress stillbirth history on insomnia at 18 and 32 weeks of gestation, and miscarriage history on insomnia at 18 weeks of gestation, assuming that insomnia in the index pregnancy could also represent the situations in previous pregnancies.

All analyses were performed using R 3.5.1 (R Foundation for Statistical Computing, Vienna, Austria). Two-sample MR analyses were conducted using the “TwoSampleMR” R package [26].

## Results

Table 1 summarizes the characteristics of included women from UKB, ALSPAC, BiB, MoBa and FinnGen. The SNP-insomnia associations in UKB and ALSPAC are listed in S4 Table (S2 File). The 81 SNPs explained approximately 0.42% variance of insomnia among the 208,140 UKB women included in this study (S4 Table in S2 File), and the mean F-statistic of the 81 SNPs was 11. The pooled 81 SNP-insomnia associations at 18 (OR 1.02 per effect allele, 95% CI: 1.01, 1.03) and 32 (OR 1.02, 95% CI: 1.01, 1.03) weeks of gestation in ALSPAC were in the same direction as (but weaker than) the pooled association in the original GWAS of UKB plus 23andMe women (OR 1.05, 95% CI: 1.05, 1.06). The SNP-outcome associations in UKB, ALSPAC, BiB and MoBa are listed in S5 Table (S2 File).

**Table 1.** Characteristics of the women in UK Biobank (UKB), Avon Longitudinal Study of Parents and Children (ALSPAC), Born in Bradford (BiB), Norwegian Mother, Father and Child Cohort Study (MoBa), and FinnGen.

| Variable <sup>a</sup> | UKB (N=208,140) | ALSPAC (N=6,826) | BiB (N=2,940) | MoBa (N=14,584) | FinnGen (N=~123,579) |
| --- | --- | --- | --- | --- | --- |
|  | *Mean (standard deviation)* |  |  |  |  |
| Maternal age at delivery (years) | 25.5 (4.6) <sup>b</sup> | 28.7(4.7) | 26.8 (6.0) | 30.0 (4.4) | Not available |
| Maternal height (cm) | 162.7 (6.2) | 164.3 (6.7) | 164.4 (6.1) | 168.3 (5.5) | Not available |
| Maternal body mass index (kg/m<sup>2</sup>) | 27.0 (5.1) | 22.9 (3.7) | 26.7 (6.0) | 24.0 (4.2) | Not available |
| Gestational age (weeks) | 38.9 (3.8) <sup>c</sup> | 39.6 (1.7) | 39.7 (1.9) | 39.6 (1.7) | Not available |
| Offspring birthweight (grams) | 3186.7 (547.6) | 3441.5 (523.0) | 3357.9 (571.2) | 3640.8 (513.4) | Not available |
|  | *N (%)* |  |  |  |  |
| Maternal education |  |  |  |  |  |
| O levels/GCSEs or equivalent and below | 91,093 (44.2) | 4,043 (59.5) | 1,400 (47.6) | 260 (1.9) | Not available |
| A levels/AS levels or equivalent | 48,059 (23.3) | 1,719 (25.3) | 485 (16.5) | 4356 (31.8) | Not available |
| College or university degree | 66,873 (32.5) | 1,035 (15.2) | 551 (18.7) | 9072 (66.3) | Not available |
| Maternal ever smoking | 85,501 (41.3) | 1,450 (21.6) <sup>d</sup> | 911 (31.0) <sup>d</sup> | 1106 (8.8) <sup>d</sup> | Not available |
| Maternal ever drinking | 191,010 (91.2) | 4,580 (70.2) <sup>d</sup> | 1,793 (61.0) <sup>d</sup> | 3644 (29.7) <sup>d</sup> | Not available |
| Offspring sex, male | Not available | 3,430 (50.2) | 1,504 (51.2) | 7412 (50.9) | Not available |
| Number with fetal genotype data | 0 | 4,625 (67.8) | 1,855 (63.1) | 12,183 (83.5) | Not available |
|  | *N cases / N controls (Prevalence, %)* |  |  |  |  |
| Stillbirth | 4907/139,034 (3.4) | 48/4546 (1.0) | 31/2588 (1.2) | 51/9998 (0.5) | Not available |
| Miscarriage | 42,717/139,034 (23.5) | 1378/4546 (23.3) | 14/2588 (0.5) | 2677/9998 (21.1) | 9113/89,340 (9.3) |
| Gestational diabetes | 726/200,536 (0.4) | 34/6283 (0.5) | 136/2657 (4.9) | 113/14,375 (0.8) | 5687/117,892 (4.6) |
| Hypertensive disorders of pregnancy | 2138/206,002 (1.0) | 1099/5698 (16.2) | 347/2159 (13.8) | 1892/12,652 (13.0) | 4255/114,735 (3.6) |
| Perinatal depression | 5178/25,130 (17.1) | 423/5896 (6.2) | 312/2245 (12.2) | 579/13,865 (4.0) | Not available |
| Preterm birth | 556/4862 (10.3) <sup>c</sup> | 285/4931 (5.5) | 172/2706 (6.0) | 495/12,846 (3.7) | 5480/98,626 (5.3) |
| Low offspring birthweight | 13,429/149,084 (8.3) | 337/6376 (5.0) | 167/2725 (5.8) | 245/13,690 (1.8) | Not available |
| High offspring birthweight | 2716/149,084 (1.8) | 113/6376 (1.7) | 42/2725 (1.5) | 621/13,690 (4.3) | Not available |

<sup>a</sup> In UKB, these variables were measured at the recruitment that is typically 31.1 years after pregnancy.
<sup>b</sup> We report maternal ages at giving their first live birth. UKB women were recruited with an average age of 56.5 (standard deviation 7.9) years.
<sup>c</sup> Gestational age was available only in a small subset of UKB women (N=7280).
<sup>d</sup> These were maternal ever smoking/drinking in pregnancy.

### Two-sample MR

In the analysis combining all cohorts, lifetime tendency to insomnia was associated with higher risks of all outcomes with point estimate OR ranging from 1.20, for GD, to 3.56 for perinatal depression (Fig 2). Despite combining data from the largest genetic studies available estimates were imprecise, with 95% CIs for all but three outcomes including the null. The three that did not include the null were miscarriage (OR 1.60, 95% CI: 1.18, 2.17), perinatal depression (OR 3.56, 95% CI: 1.49, 8.54) and LBW (OR 3.17, 95% CI: 1.69, 5.96). S2 Fig (in S1 File) shows IVW results for leave-one (study)-out analysis. Results were broadly consistent, with the point estimates showing some differences with stillbirth, GD and PTB, though CIs were very wide for some outcomes.

**Fig 2.**
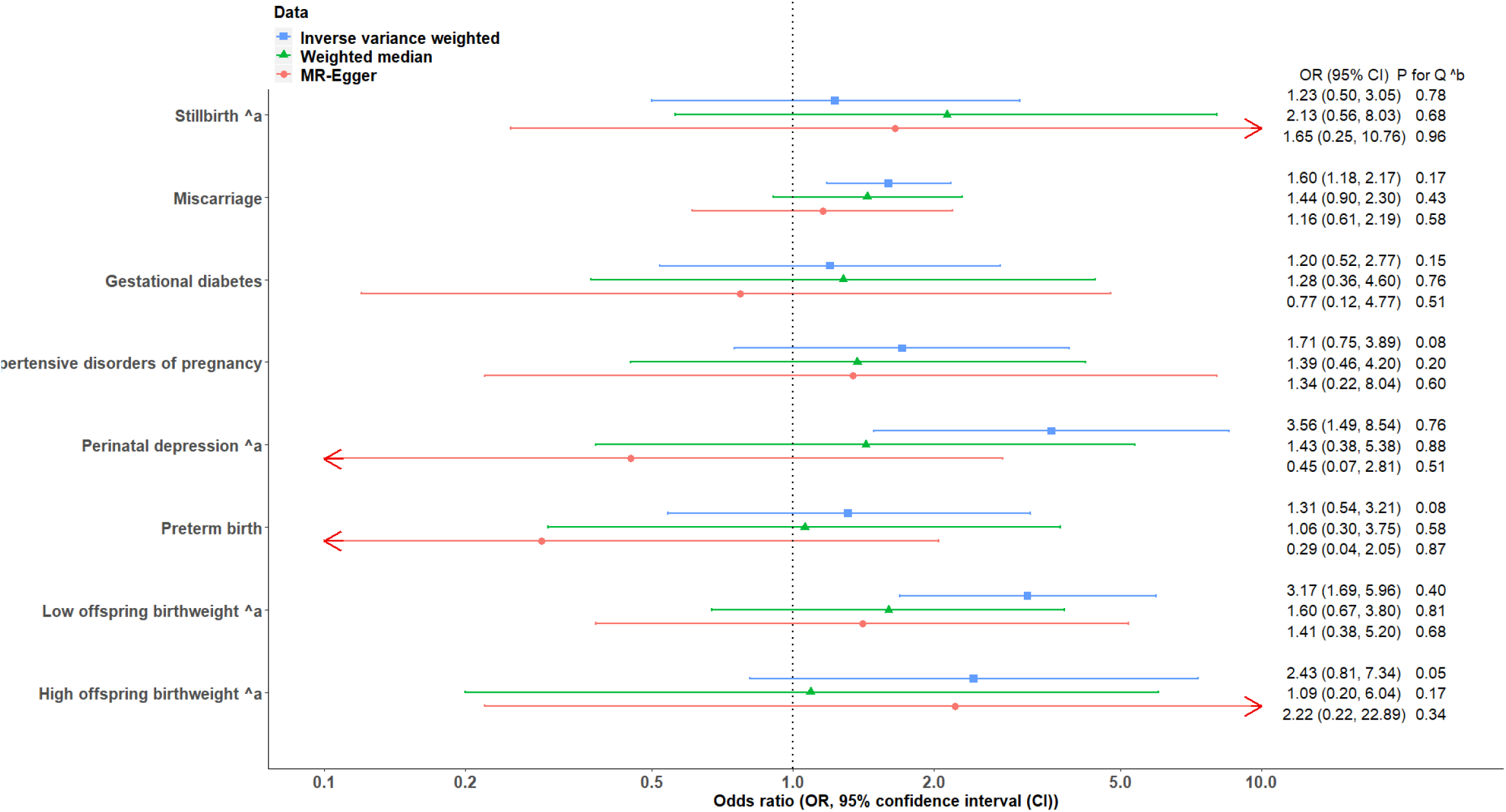
Two-sample Mendelian randomization (MR) estimates for causal effects of insomnia on adverse pregnancy and perinatal outcomes, meta-analysing UK Biobank, the birth cohorts, and FinnGen. ^a^ For those outcomes that FinnGen did not contribute to. ^b^ P-value for Cochran’s Q-statistic <0.05 suggests between-study heterogeneity in the meta-analysis.

Sensitivity analyses using weighted median and MR-Egger for all outcomes were directionally consistent, with the exception of MR-Egger results for GD, perinatal depression and PTB (Fig 2). Between-SNP heterogeneity for MR analyses was observed with LBW and HDP (S6 Table in S2 File), but leave-one (SNP)-out analyses were consistent with the main IVW estimates including all SNPs for all outcomes (S3-5 Figs in S1 File). The MR-Egger intercept p-value indicated unbalanced horizontal pleiotropy only for perinatal depression in UKB (S6 Table in S2 File). Adjusting for fetal genotype (only possible in the birth cohorts) did not alter the SNP-outcome associations with stillbirth, miscarriage, LBW or HBW; SNP-outcome associations with GD, HDP and perinatal depression were slightly attenuated; SNP-PTB associations moved slightly away from the null (S6 Fig in S1 File).

### Multivariable regression in ALSPAC

S7 Table (S2 File) summarizes the characteristics of women from ALSPAC who contributed to these analyses. After adjusting for potential confounders, there were associations of insomnia at 18 and 32 weeks of gestation with stillbirth history, miscarriage history (only assessed for 18 weeks), GD (32 weeks only), HDP and perinatal depression, with similar magnitudes of association to those seen for the MR analyses, and with imprecision meaning some 95% CIs included the null (Fig 3). Associations with stillbirth history (OR for insomnia at 18 weeks 2.10, 95% CI: 1.20, 3.69), miscarriage history (OR for insomnia at 18 weeks 1.30, 95% CI: 1.12, 1.51), and perinatal depression (OR for insomnia at 18 weeks 2.96, 95% CI: 2.42, 3.63), were the most reliable with CIs that did not include the null (Fig 3).

**Fig 3.**
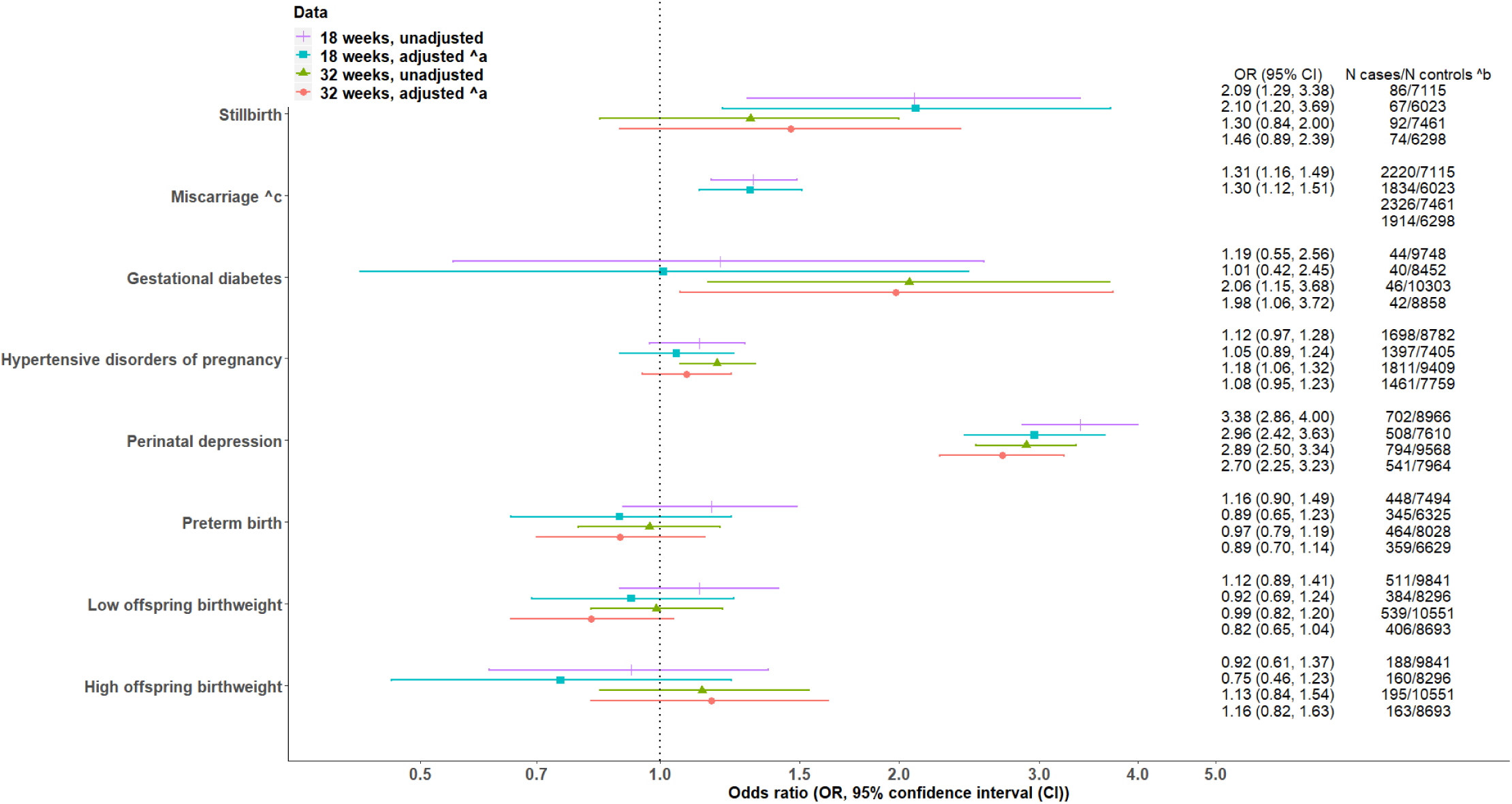
Multivariable regression associations of insomnia at 18 and 32 weeks of gestation with adverse pregnancy and perinatal outcomes in Avon Longitudinal Study of Parents and Children (ALSPAC). ^a^ We adjusted for maternal age at time of delivery, education, body mass index at 12 weeks of gestation, smoking status in pregnancy and alcohol intake in the first three month of ALSPAC pregnancy, and household occupational social class. ^b^ The numbers of women in adjusted models are slightly smaller than those in crude models due to missingness (<8%) in these covariates. ^c^ We only report results for insomnia at 18 weeks for miscarriage as by definition that has to occur early in pregnancy (i.e. prior to 20-24 weeks of gestation in the different populations included in the analyses).

## Discussion

To our knowledge this is the first MR study to explore the relationship of insomnia with pregnancy and perinatal outcomes. We interpreted the MR results as reflecting a lifetime tendency to insomnia on the basis that SNPs are determined at conception, and evidence suggested that with similar analyses of other exposures (e.g. blood pressure and C-reactive protein) this is the case [36, 37]. We interpreted the multivariable regression results as reflecting associations of insomnia during pregnancy, though we could not distinguish this from pre-existing insomnia as we did not have information on sleep traits before conception. The associations of the insomnia genetic IVs with reported insomnia during pregnancy in ALSPAC provided some support that the exposures in our MR and multivariable regression analyses had some consistency with each other. Overall, our MR results provide evidence that a lifetime tendency to insomnia may increase the risk of stillbirth, miscarriage, GD, HDP, perinatal depression, PTB, LBW and HBW, with ORs of potential clinical importance (range 1.20 to 3.56) for all of these. However, we acknowledge that MR analyses are statistically inefficient and despite combining all potentially relevant studies in order to increase the sample size, results were imprecise with all but three outcomes having 95% CI that included the null. Thus, we have strongest MR evidence for insomnia increasing the risk of miscarriage, perinatal depression, and LBW. Results for miscarriage and perinatal depression are replicated in multivariable regression analyses,but with the exception of results for LBW, which were much closer to the null.

Our findings in both MR and multivariable regression of increased risks of perinatal depression are consistent with the systematic review and meta-analysis of observational studies [7], and with RCTs suggesting that pregnancy interventions to reduce insomnia decrease perinatal depression [11, 12]. Whilst previous studies have shown that pregnancy supine sleeping position (which is associated with insomnia) to be associated with stillbirth [6, 9], recent systematic reviews have only identified one cross-sectional study (N=222) of the association of insomnia with stillbirth [6, 8], and we did not identify any previous studies of insomnia associations with miscarriage. Thus, our novel findings of a possible effect of insomnia on miscarriage in both MR and multivariable regression analyses warrants replication. Previous systematic reviews of observational associations of insomnia with GD, HDP and PTB have some consistency with our main MR results, but few observational studies investigated LBW or HBW [6, 8]. For example, ORs from the meta-analyses for GD (1.37 combining 8 studies vs 1.20 in MR), HDP (1.72 combining 2 studies vs 1.71 in MR), PTB (1.49 combining 8 studies vs 1.31 in MR) were very similar to our MR analyses [6], despite the differing definitions of insomnia and outcomes and different assumptions for MR and multivariable regression [38].

Several mechanisms have been suggested for why insomnia might influence pregnancy and perinatal outcomes, including insomnia resulting in increased risks of adiposity, insulin resistance and other cardiometabolic outcomes that could then influence related pregnancy outcomes (GD, HDP, LBW and HBW) and influence placentation and hence miscarriage, stillbirth and PTB. MR analyses support effects of insomnia on coronary heart disease, higher glycated haemoglobin, and higher glycoprotein acetyls (an inflammatory marker) in general populations of women and men [20, 39, 40]. Thus, an increase in cardio-metabolic risk and inflammation may mediate effects of insomnia on miscarriage and LBW, and outcomes for which our MR analyses are currently imprecise. Similarly, MR analyses have found a potential effect of insomnia on depressive symptoms [20], which is coherent with our findings in relation to perinatal depression.

### Study strengths and limitations

Key strengths of our study are that (I) it is the first study to use MR to explore potential effects of insomnia on pregnancy and perinatal outcomes; (II) we compared those MR findings with multivariable regression results of insomnia symptoms in pregnancy, adjusting for a *priori* defined key confounders; (III) we explored a range of pregnancy and perinatal outcomes. To our knowledge, this is the first study to explore associations with miscarriage and stillbirth, using either multivariable regression or MR.

Our MR analyses may be biased by horizontal pleiotropy, particularly given our previous research showing that SNPs for insomnia are also associated with several factors that could influence pregnancy and perinatal outcomes, including education, age at first live birth, and smoking [33]. We explored this potential with a range of sensitivity analyses, including exploring between-SNP heterogeneity and using weighted median and MR-Egger methods that are more robust to such bias than IVW [30]. Results from these sensitivity analyses were broadly consistent with our main analyses but were less precise (i.e. had wider CIs). Adjusting for fetal genotype did not alter results suggesting that bias due to fetal genotypic effects is unlikely. We did not further adjust for paternal genotype because of limited data with paternal, maternal and offspring genotype. Furthermore, the most plausible mechanism for paternal genotype to affect pregnancy outcomes is via fetal genotype, which we have adjusted for.

Interpretation of the effect estimates from our MR analyses requires a further assumption of monotonicity in the SNP-insomnia associations. This requires that all of the women with genetic IVs related to higher liability to insomnia symptoms should report more symptoms (compared to those with fewer alleles related to insomnia) – i.e. that they are ‘compliers’ [41]. Whilst we cannot test this assumption, the similarity of our MR and multivariable regression estimates for miscarriage and perinatal depression, provides some evidence that it may not have been violated for these outcomes.

Both our MR and multivariable regression estimates could be vulnerable to selection bias, which has been extensively discussed in previous papers [42-44]. Specific to UKB, it is a highly selective sample that is healthier and better educated than the general UK adult population [45]. Moreover, information on perinatal depression and PTB was only available in a subsample of UKB women and such missingness might not be at random [46, 47]. By definition our study only includes women who have experienced at least one pregnancy, and if insomnia influences fertility then our results might be biased [48]. However, we are not aware of robust evidence of insomnia (or SNPs related to insomnia) influencing infertility or number of children [49, 50], suggesting any selection bias through only including pregnant women is unlikely to have a meaningful impact on our MR estimates [48, 51].

Insomnia was measured via one self-administrated question in both UKB and ALSPAC, which could mean the binary exposure is misclassified. Non-differential misclassification of insomnia would be expected to bias MR results away from the null (given the attenuated genetic IVs – insomnia associations is the denominator), but multivariable regression results towards the null [52, 53]. Similarly, there may be misclassification in some of our outcomes because of the absence of universal testing (e.g. GD in ALSPAC [54]), assessment via self-report questionnaires (e.g. BW in UKB) or differences between studies in definitions (e.g. in older women in UKB the gestational age thresholds for defining stillbirth and miscarriage would have differed from those used in the more contemporary birth cohorts). Non-differential misclassification of our binary outcomes would be expected to bias both MR and multivariable regression results towards the null [52, 53]. Moreover, the first live-born babies of UKB women are known to be lighter than babies with various birth orders from the more contemporary birth cohorts [55, 56].

Both MR and multivariable regression analyses of miscarriage and stillbirth assessed the associations of ever experiencing either of them mainly up to the point of recruitment. For the multivariable regression analysis in ALSPAC, insomnia was reported after the outcomes had occurred and it is possible that previous miscarriage or stillbirth might have influenced sleep in the index pregnancy (where insomnia was reported), e.g. due to anxiety [3]. The similarity of the multivariable regression and MR results for miscarriage, stillbirth, GD, HDP and perinatal depression suggests residual confounding is unlikely to have biased regression results for these outcomes. The attenuated to the null associations for preterm birth, LBW and HBW compared to MR results suggests possible masked confounding or other biases specifically affecting these but not other outcomes. Despite having a large sample size and our multivariable analyses being larger than most previous studies, several of our MR and multivariable regression estimates are imprecise. Our study is limited to women of European ancestry, and we cannot assume that our results generalize to other populations.

In conclusion, our study raises the possibility of adverse causal effects of insomnia on miscarriage, perinatal depression, and LBW. Interventions to improve healthy sleep in women of reproductive age might be beneficial to a healthy pregnancy. However, we acknowledge the need for further MR studies based on larger GWAS of pregnancy and perinatal outcomes, larger observational studies, and studies in women from ethnic backgrounds other than White European.

## Data Availability

We used both individual participant cohort data and publicly available summary statistics. We present summary statistics that we generated from those individual participant cohort data in S4 & 5 Tables. Full information on how to access UKB data can be found at its website (https://www.ukbiobank.ac.uk/researchers/). All ALSPAC data are available to scientists on request to the ALSPAC Executive via this website (http://www.bristol.ac.uk/alspac/researchers/), which also provides full details and distributions of the ALSPAC study variables. Similarly, data from BiB are available on request to the BiB Executive (https://borninbradford.nhs.uk/research/how-to-access-data/). Data from MoBa are available from the Norwegian Institute of Public Health after application to the MoBa Scientific Management Group (see its website https://www.fhi.no/en/op/data-access-from-health-registries-health-studies-and-biobanks/data-access/applying-for-access-to-data/ for details). Summary statistics from FinnGen are publicly available on its website (https://finngen.gitbook.io/documentation/data-download).

## Acknowledgments

This research has been conducted using the UKB Resources under application number 23938. The authors would like to thank the participants and researchers from UKB who contributed or collected data. We are extremely grateful to all the families who took part in this study, the midwives for their help in recruiting them, and the whole ALSPAC team, which includes interviewers, computer and laboratory technicians, clerical workers, research scientists, volunteers, managers, receptionists and nurses. BiB is only possible because of the enthusiasm and commitment of the Children and Parents in BiB. We are grateful to all the participants, teachers, school staff, health professionals and researchers who have made BiB happen. This research has been conducted using MoBa data using application number 2552. MoBa is supported by the Norwegian Ministry of Health and Care services and the Ministry of Education and Research. We are grateful to all the participating families in Norway who take part in this on-going cohort study. We thank the Norwegian Institute of Public Health (NIPH) for generating high-quality genomic data. This research is part of the HARVEST collaboration, supported by the Research Council of Norway (#229624). We also thank the NORMENT Centre for providing genotype data, funded by the Research Council of Norway (#223273), South East Norway Health Authority and KG Jebsen Stiftelsen. We further thank the Center for Diabetes Research, the University of Bergen for providing genotype data and performing quality control and imputation of the data funded by the ERC AdG project SELECTionPREDISPOSED, Stiftelsen Kristian Gerhard Jebsen, Trond Mohn Foundation, the Research Council of Norway, the Novo Nordisk Foundation, the University of Bergen, and the Western Norway health Authorities (Helse Vest). The authors thank FinnGen investigators for sharing their summary-level data.

