## Supplementary material for "Associations of insomnia on pregnancy and perinatal outcomes: Findings from Mendelian randomization and conventional observational studies in up to 356,069 women": S1 File

**Supplementary text and figures**

### S1 Text. Descriptions of each cohort

#### *UK Biobank (UKB)*

All people in the UK National Health Service registry aged between 40-69 years and living within a 25 mile radius from one of 22 study centres were invited to participate between 2006-2010 [1]. In total 503,325 adults (5.5% of the ~9.2 million invited) were recruited into UKB [1]. Participants who had a valid email address (N=339,229) were invited to fill in a detailed online questionnaire assessing their mental health after January 2015, and 158,835 participants fully completed it by October 2017 [2]. Ethical approval for UKB was obtained from the North West Multi-centre Research Ethics Committee, and our study was performed under UKB application number 23938.

Genotyping, pre-imputation quality control, and imputation procedures were described in detail elsewhere [3], and are briefly summarized here. UKB men and women were genotyped on two arrays. The first ~50,000 samples were genotyped on the UK BiLEVE array and the remaining ~450000 samples were genotyped on the UK Biobank Axiom array. Genotype data were imputed against two references panels: Haplotype Reference Consortium (HRC) panel and UK10K + 1000 Genomes panel. We used the imputed data released by UKB in March 2018, and applied in-house post-imputation quality controls (QC, i.e. genetic sex same as reported sex, XX or XY in sex chromosome and no outliers in heterozygosity and missing rates) [4]. Women of European descent with qualified genotype data were eligible for inclusion in our Mendelian randomization (MR) analyses (N=208,140, see S1 Fig A).

Information about observed confounders were collected at the initial assessment. Age (years) was derived based on date of birth and date of attending an initial assessment centre. Women were asked to report their age when they had their only one child, or the first child if they had given birth to more than one child. Anthropometric measures included standing height (cm) and body mass index (BMI, kg/m$^2$, constructed from standing height and weight). Participants were also asked to report (1) the qualifications they achieved, and we grouped their highest qualifications into three levels: O levels/GCSEs or equivalent and below, A levels/AS levels or equivalent, and College or university degree; (2) their smoking status (current/former/never), and we derived a binary ever versus never measure of smoking status by combining current and former smokers; (3) frequency of drinking alcohol at six levels from ‘never’ to ‘daily or almost daily’, and we derived a binary variable for the comparison between cohorts.

#### *Avon Longitudinal Study of Parents and Children (ALSPAC)*

Pregnant women resident in Avon, UK with expected dates of delivery 1$^{st}$ April 1991 to 31$^{st}$ December 1992 were invited to take part in the study [5, 6]. The initial number of pregnancies enrolled is 14,541 (for these at least one questionnaire has been returned or a ‘Children in Focus’ clinic data had been attended by 19/07/99) [5, 6]. Of these initial pregnancies, there was a total of 14,676 foetuses, resulting in 14,062 live births and 13,988 children who were alive at 1 year of age [5, 6]. Our study relied on 13,867 pregnancies from a total of 13,761 women, and most pregnancies were recruited at the first antenatal clinic visit in the first trimester of pregnancy [5]. Questionnaires were sent at regular intervals during pregnancy, and biological samples were taken from parents and children including blood samples, from which DNA was extracted. The study website contains details of all the data that is available through a fully searchable data dictionary and variable search tool (http://www.bristol.ac.uk/alspac/researchers/our-data). Ethical approval for the study was obtained from the ALSPAC Ethics and Law Committee and the Local Research Ethics Committees. Consent for biological samples has been collected in accordance with the Human Tissue Act (2004). Informed consent for the use of data collected via questionnaires and clinics was obtained from participants following the recommendation of the ALSPAC Ethics and Law Committee at the time.

Genotyping, pre-imputation quality control, and imputation procedures were described in detail elsewhere [7], and are briefly summarized here. ALSPAC mothers were genotyped using Illumina human660K quad SNP chip, and ALSPAC children were genotyped using Illumina HumanHap550 quad genome-wide SNP genotyping platform. Genotype data for both ALSPAC mothers and children were imputed against HRC v1.1 reference panel, after a similar QC procedure (minor allele frequency (MAF) ≥1%, a call rate ≥95%, in Hardy-Weinberg equilibrium (HWE), correct sex assignment, no evidence of cryptic relatedness, and of European descent). Women of European descent with qualified genotype data and live-born singleton offspring were eligible for inclusion in our MR analyses (N=6826, see S1 Fig. B).

Maternal age at delivery (years) was derived from date of delivery and date of birth. Maternal height and pre-pregnancy weight were self-reported at 12 weeks of gestation and were used to calculate BMI. Mothers were asked to report their highest educational qualification, and we categorized the answers into three levels ("CSE/Vocational/O level", "A level" and "Degree"). We used self-reported tobacco smoking in the first three months of pregnancy and in the last two weeks at 18 weeks of gestation to construct a binary ever versus never smoked in pregnancy. Never smokers were women who did not smoke in both time periods, while ever smokers were women who smoked in either of them. Mothers reported their frequency of alcohol use in the first three months and in the last two months of pregnancy at six levels, and we also derived a binary ever versus never measure of alcohol consumption. Sex of offspring (ALSPAC children) was recorded. Both maternal and paternal occupational social class were reported by ALSPAC mothers at 32 weeks of gestation. They were defined using the UK Registrar General's occupational coding, and were coded from social class V (unskilled manual) to social class I (professional) [8]. We generated the household occupational social class based on whichever was the higher one.

***Born in Bradford (BiB)***

BiB is a birth cohort that recruited 13,776 pregnancies to 12,453 women resident in the Bradford metropolitan district (a city in the North of England), with expected dates of delivery between 2007-2010 [9]. Most pregnancies were recruited during an oral glucose tolerance test undertaken between 24-28 weeks of gestation, and to which all pregnant women in Bradford were invited [9]. BiB reflects the multicultural mix profile of Bradford, with approximately 50% of women being of South Asian descent [9]. The study website (https://borninbradford.nhs.uk/) provides details of all available data. Ethical approval for BiB was obtained from the Bradford Research Ethics Committee.

Genotyping, pre-imputation quality control, and imputation procedures were described elsewhere [10], and we briefly summarize here. Both BiB mothers and BiB children were genotyped using Illumina HumanCoreExome chip. Genotype data for both of them were imputed against UK10K + 1000 Genomes reference panel, after a similar QC procedure (a call rate ≥99.5%, correct sex assignment, no evidence of cryptic relatedness, correct ethnicity assignment). Two subsets of individuals were declared based on a combination of principal component analysis and self-reported ethnicity. To combine the MR estimates with those in other cohorts, only women of European descent with qualified genotype data and live-born singleton offspring were eligible for inclusion in our analyses (N=2940, see S1 Fig C).

Maternal age (years) was recorded when participants received their oral glucose tolerance test (OGTT) at recruitment. Maternal height and weight were recorded at recruitment, and the highest educational qualification was collected via a questionnaire at baseline. The BiB research team derived maternal BMI (using an early pregnancy weight) and categorized their highest educational qualification into 7 groups ("<5 GCSE equivalent", "5 GCSE equivalent", "A-level equivalent", "Higher than A-level", "Other", "Don't know" and "Foreign unknown"). In the baseline questionnaire, mothers were asked to report the number of cigarettes they smoked in the first three months of pregnancy and since the fourth month of pregnancy, and a binary ever versus never measure of smoking status in pregnancy was derived. Mothers were also asked to report the frequency of drinking alcohol 3 months before pregnancy, in the first three months of pregnancy and since the fourth month of pregnancy at four levels ("Yes, once a week", "Yes, occasionally", "Yes, not specified" and "No"), from which a binary measure of alcohol consumption was derived. Sex of offspring was obtained from the Eclipse electronic maternity record.

***The Norwegian Mother, Father and Child Cohort Study (MoBa)***

MoBa is a population-based pregnancy cohort study conducted by the Norwegian Institute of Public Health. Participants were recruited from all over Norway from 1999-2008 [11]. The women consented to participation in 41% of the pregnancies. The cohort now includes 114,500 children, 95,200 mothers and 75,200 fathers [11]. The current study is based on version 12 of the quality-assured data files released for research on "Prenatal environmental exposures and pregnancy outcomes-Mendelian randomization analysis". The establishment of MoBa and initial data collection was based on a license from the Norwegian Data Protection Agency and approval from The Regional Committees for Medical and Health Research Ethics. The MoBa cohort is now based on regulations related to the Norwegian Health Registry Act. The current study was approved by The Regional Committees for Medical and Health Research Ethics. The Medical Birth Registry (MBRN) is a national health registry containing information about all births in Norway. MoBa has been linked to the Medical Birth Register of Norway (MBRN, established in 1967), using unique personal identification numbers [11].

Blood samples were obtained from both parents during pregnancy and from mothers and children (umbilical cord) at birth [12]. Genotyping, pre-imputation quality control, and imputation procedures were described in detail elsewhere [13, 14], and we briefly summarized here. There were five projects that contributed to MoBa genetics 1.0 [15], and we had to use an earlier version consisting of two projects – HARVEST and ROTTERDAM1 given their complete QC procedure. In HARVEST, MoBa mothers and children were genotyped using either Illumina HumanCoreExome12v1.1 or Illumina HumanCoreExome24v1.0. In ROTTERDAM1, MoBa mothers and children were genotyped using Illumina GSAMDv1.0. Genotype data from all batches were imputed against HRC v1.1 reference panel, after a similar QC procedure (MAF ≥5%, a call rate ≥99.2%, in HWE, correct sex assignment and no evidence of cryptic relatedness). Women of European descent with qualified genotype data and live-born singleton offspring were eligible for inclusion in our analyses (N=14,584, see S1 Fig. D).

Maternal age at delivery was recorded in MBRN, and five women were grouped as "less than 17 years". Therefore, we recoded them as "NA". Maternal weight (when they became pregnant), height and the highest education attainment were self-reported at 15 weeks of gestation. We excluded implausibly extreme values (weight less than 30 kg or more than 200 kg, height shorter than 135 cm), and then constructed maternal BMI. We grouped maternal educational attainment into three levels (9-year secondary school, high school, college or university degree) to make it comparable to that in our British cohorts. Maternal smoking in pregnancy was self-reported at both 15 and 30 weeks of gestation. For each time point, we derived a binary ever versus never measure of smoking status by combining "daily" and "sometimes" smoking, and ever smokers in pregnancy were participants who smoked at either 15 or 30 weeks of gestation, while never smokers were those who did not smoke at both time points. Maternal frequencies of alcohol consumption during 0-12, 13-24 and 25-30 weeks of gestation were self-reported at 30 weeks of gestation, with seven levels from "never" to "roughly 6-7 times a week", and we also derived a binary ever versus never measure of alcohol consumption during 0-30 weeks of gestation. Sex of offspring was also recorded in MBRN, and where records showed “not specified” or “uncertain” we recoded them as “NA”.

***FinnGen***

FinnGen is the national wide network of Finnish biobanks, including Auria Biobank, Biobank Borealis of Northern Finland, Biobank of Eastern Finland, Central Finland Biobank, Finnish Red Cross Blood Service Biobank, Finnish Clinical Biobank Tampere, Helsinki Biobank, Terveystalo Biobank, and THL Biobank [16]. Those biobanks were linked to national registries, including Drug purchase and Drug Reimbursement, Digital and Population Data Services Agency, Statistics Finland, Register of primary health care visits: AVOHILMO, Care Register for Health Care: HILMO, and Finnish cancer registry. Clinical endpoints were defined based on International Classification of Diseases (ICD)-10, and the equivalent in ICD-8 and ICD-9. The Coordinating Ethics Committee of the Helsinki and Uusimaa Hospital District has approved the FinnGen consortium (Nr HUS/990/2017), and the ethical approval of each individual study has been described in detail elsewhere [17]. FinnGen participants were genotyped with Illumina and Affymetrix chip arrays. Genotype data was imputed against SISu v3 reference panel (http://sisuproject.fi), after a QC procedure (minor allele count ≥3, a call rate ≥98%, in HWE no outliers in heterozygosity, correct sex assignment, and of Finnish ancestry). The FinnGen team released summary-level data of genome-wide association studies of more than 4000 clinical endpoints (https://www.finngen.fi/en/researchers/clinical-endpoints) at R5 wave (N=218,792 men and women).

**S1 Fig. Flow chart of each cohort**

*a. UK Biobank (UKB)*

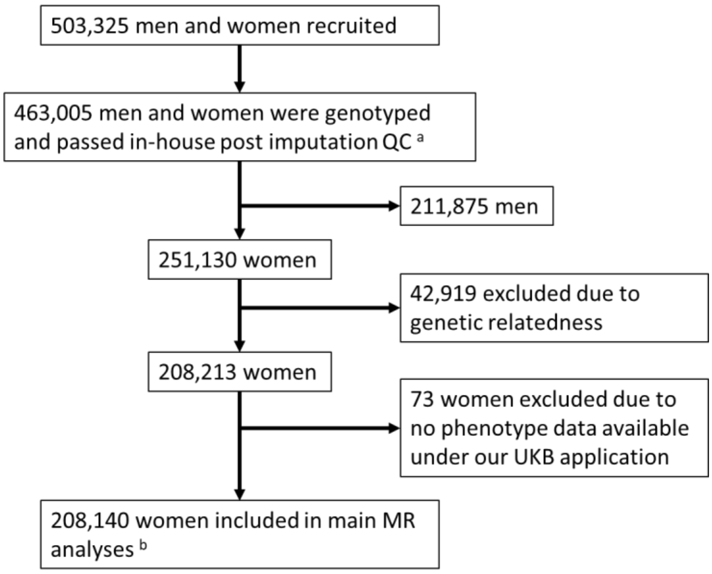


*b. Avon Longitudinal Study of Parents and Children (ALSPAC)*

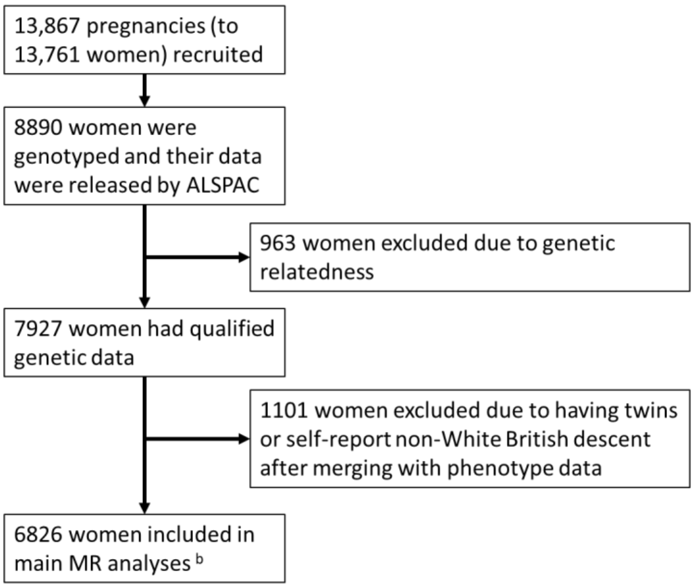

### *c. Born in Bradford (BiB)*

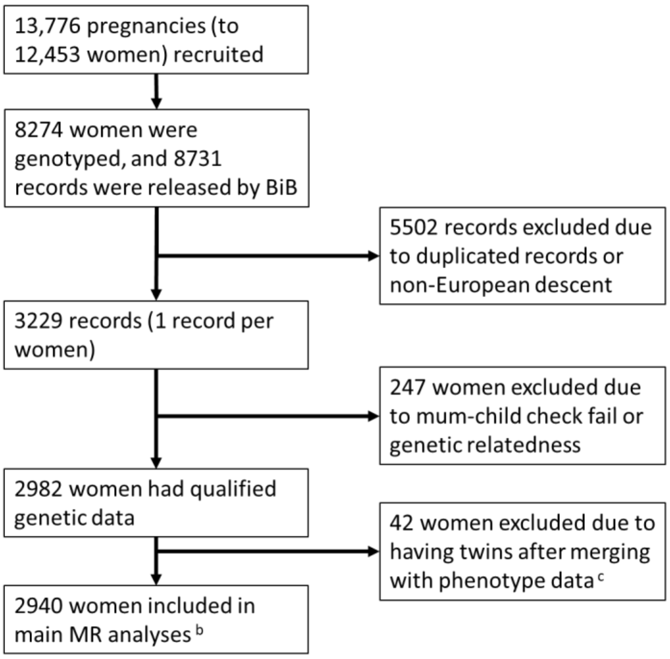


### *d. The Norwegian Mother, Father and Child Cohort Study*

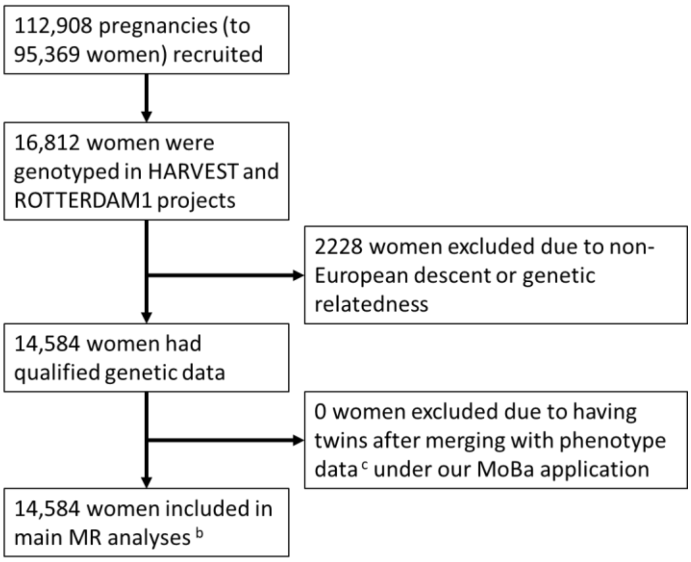


[a] We used the imputed data released by UKB in March 2018, and applied in-house post-imputation QC [4].
[b] The numbers of cases and controls for each pregnancy and perinatal outcomes are listed in Table 1.
[c] We randomly selected one pregnancy per woman in the phenotype data if multiple pregnancies were recorded.
Abbreviation: Mendelian randomization, MR; Quality control, QC.

**S2 Fig. Leave-one (study)-out analyses for causal effects of insomnia on adverse pregnancy and perinatal outcomes in Mendelian randomization using inverse variance weighted estimates.**

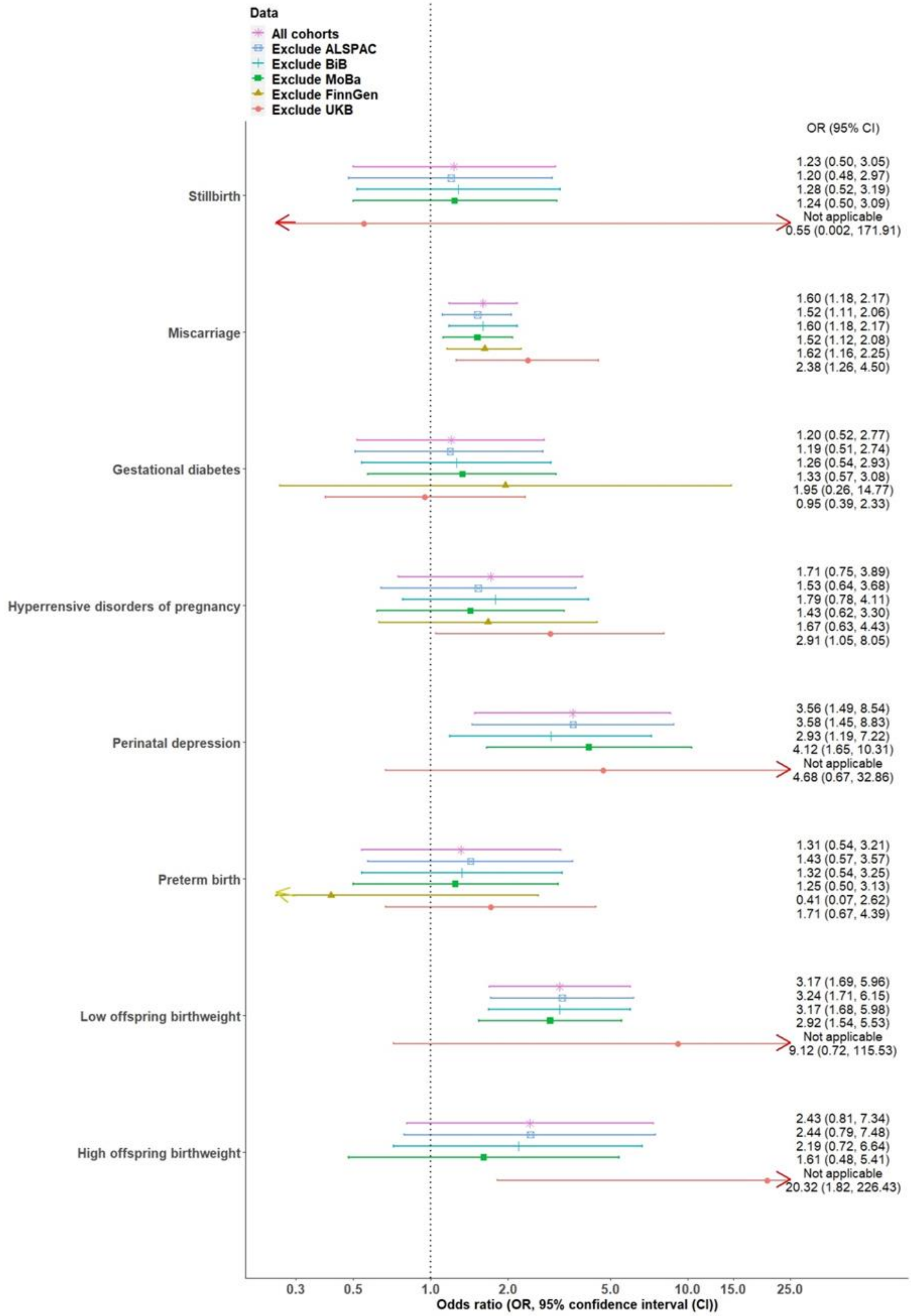


Abbreviations: ALSPAC, Avon Longitudinal Study of Parents and Children; BiB, Born in Bradford; MoBa, The Norwegian Mother, Father and Child Cohort Study; UKB, UK Biobank

**S3 Fig. Leave-one (single nucleotide polymorphisms)-out sensitivity analysis for insomnia on pregnancy and perinatal outcomes in UK Biobank (dataset A on dataset B)**

(a) Stillbirth

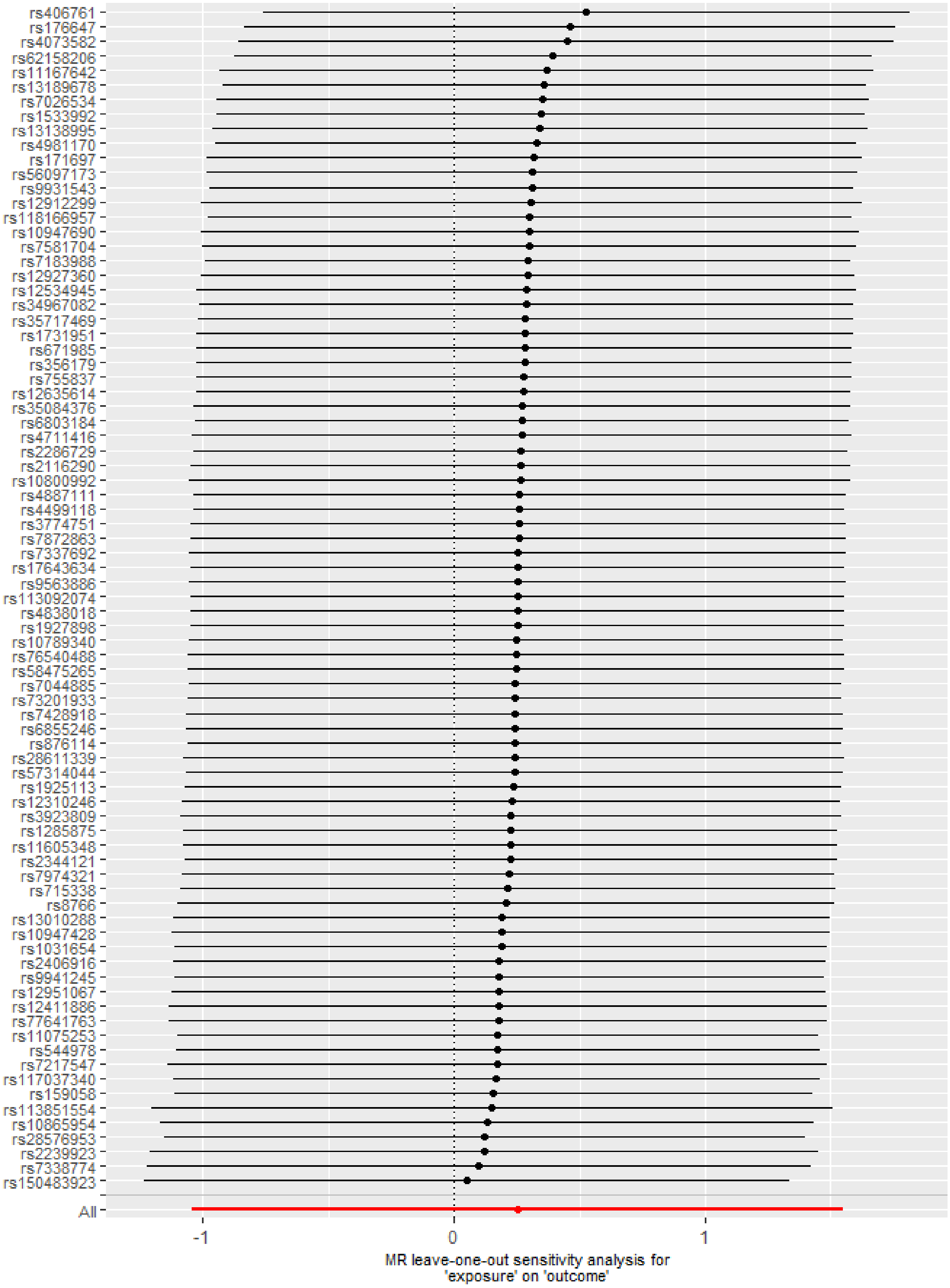

(b) Miscarriage

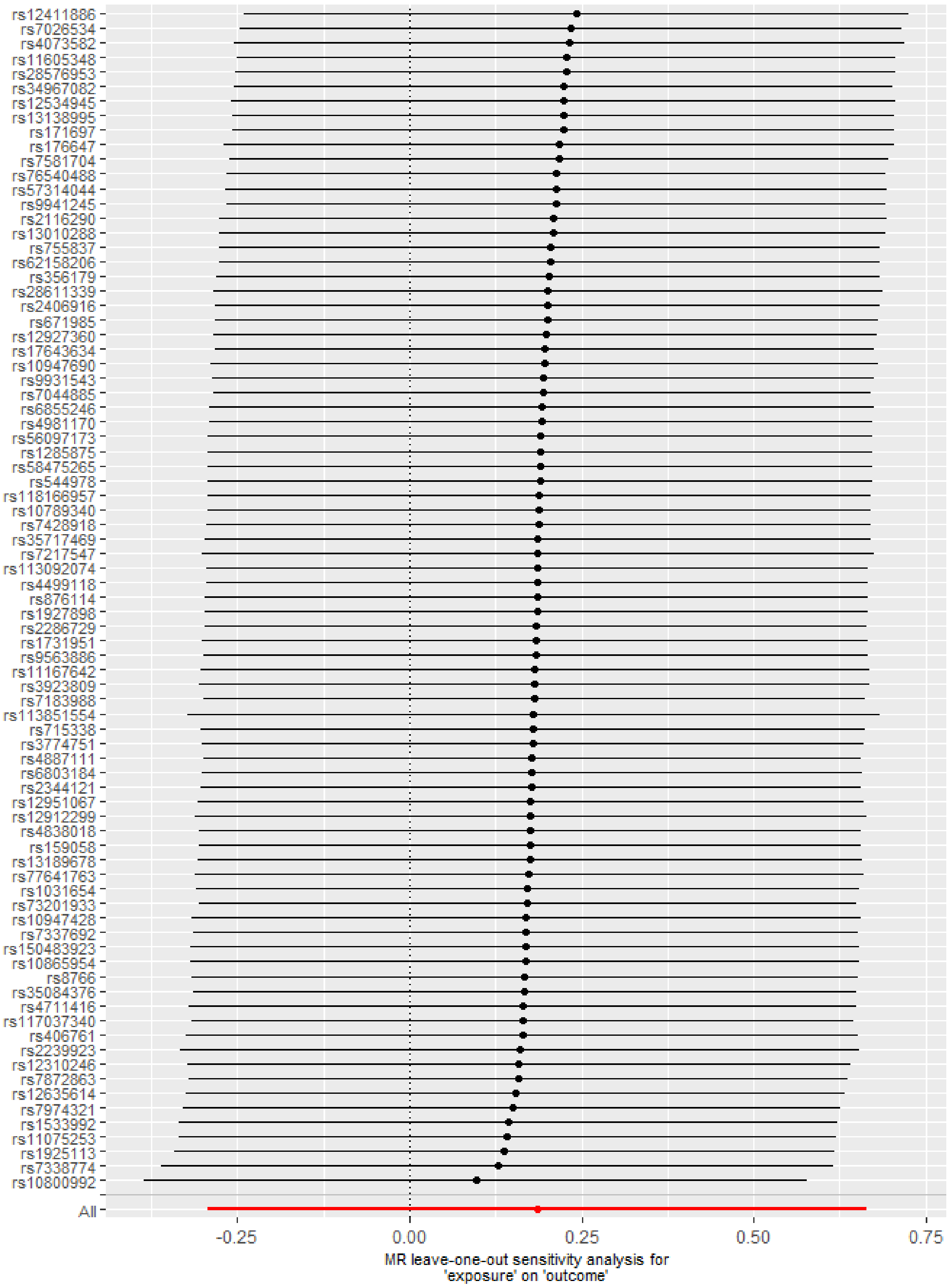

(c) Gestational diabetes

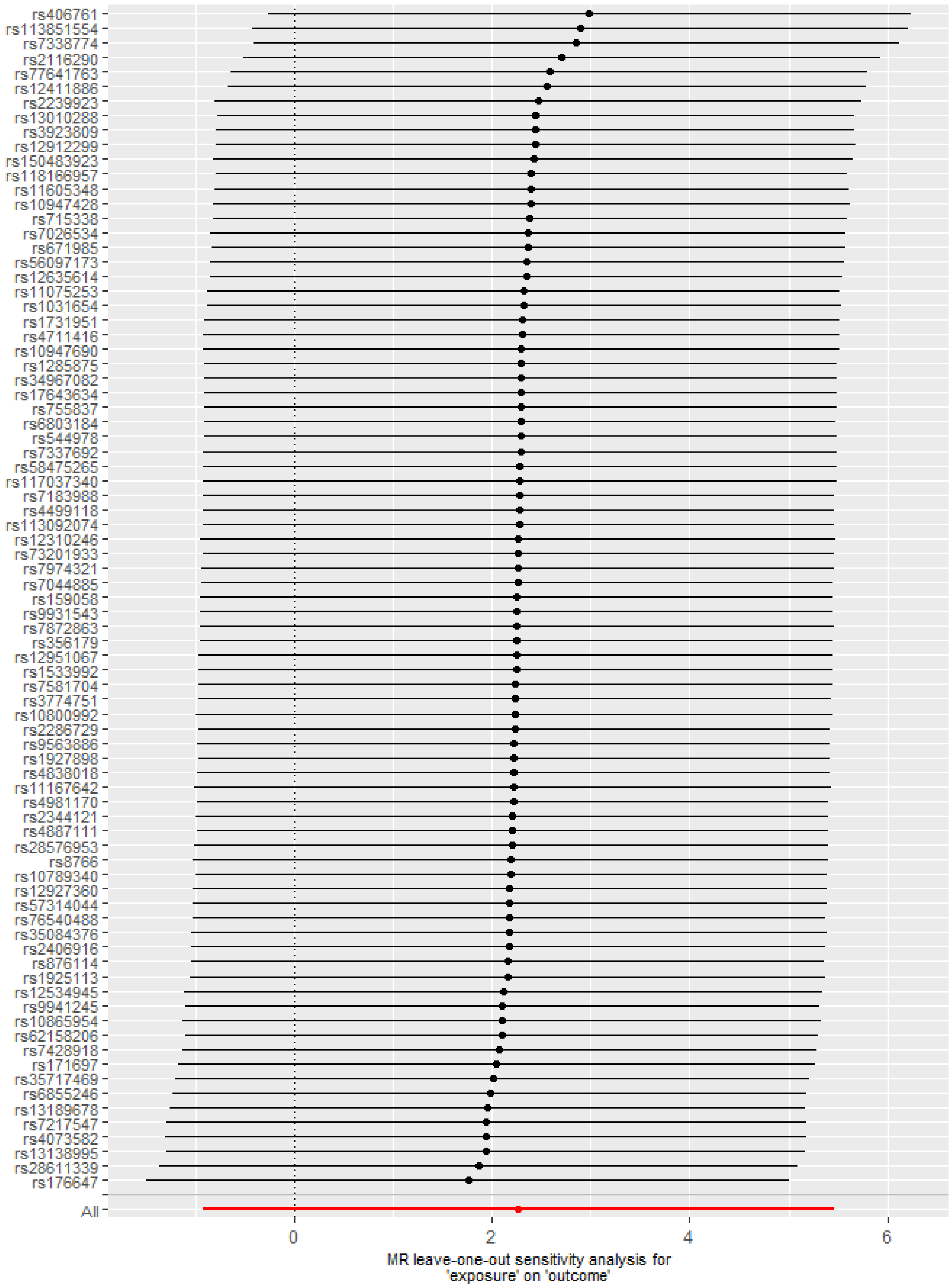

(d) Hypertensive disorders of pregnancy

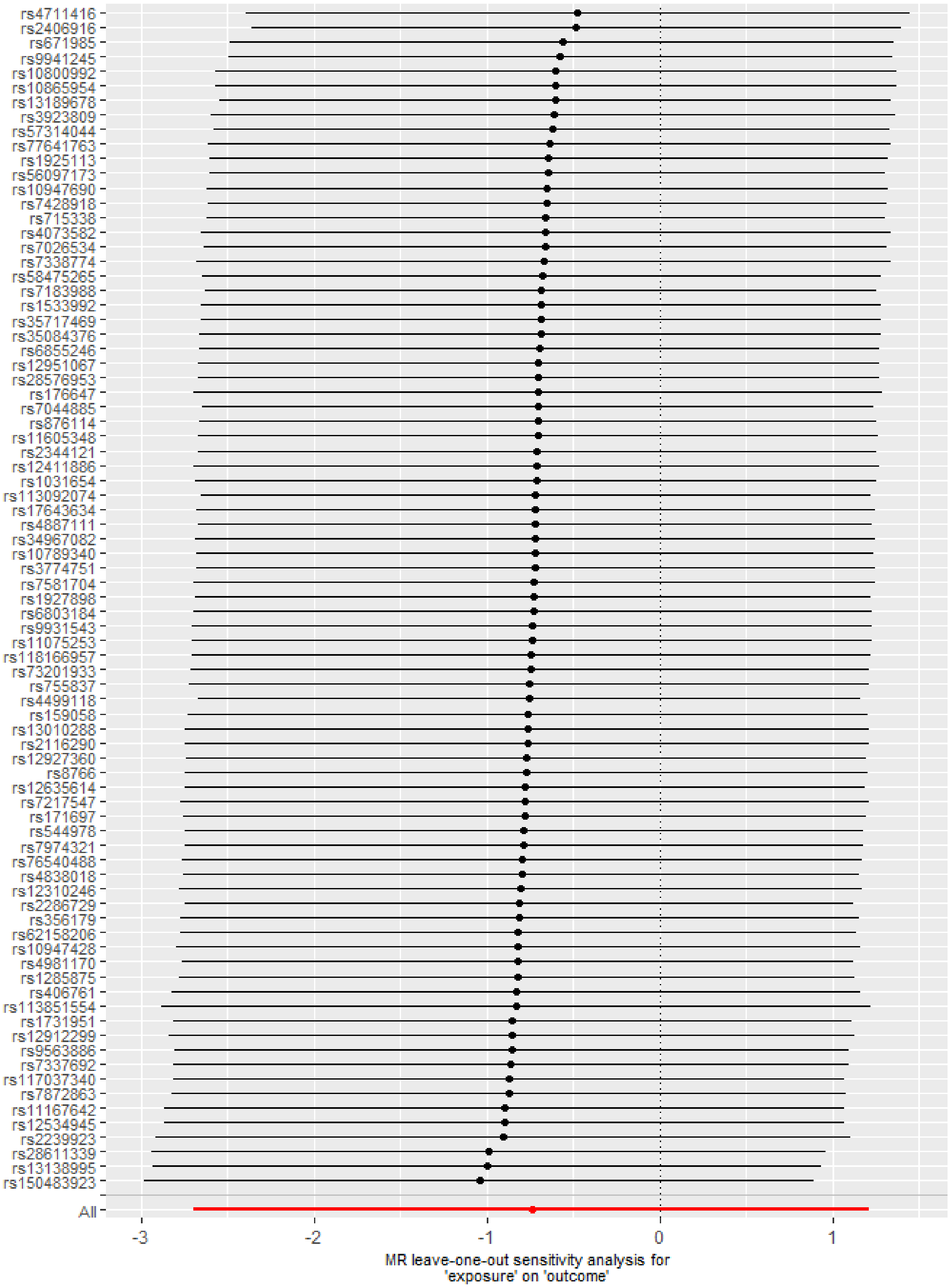

(e) Perinatal depression

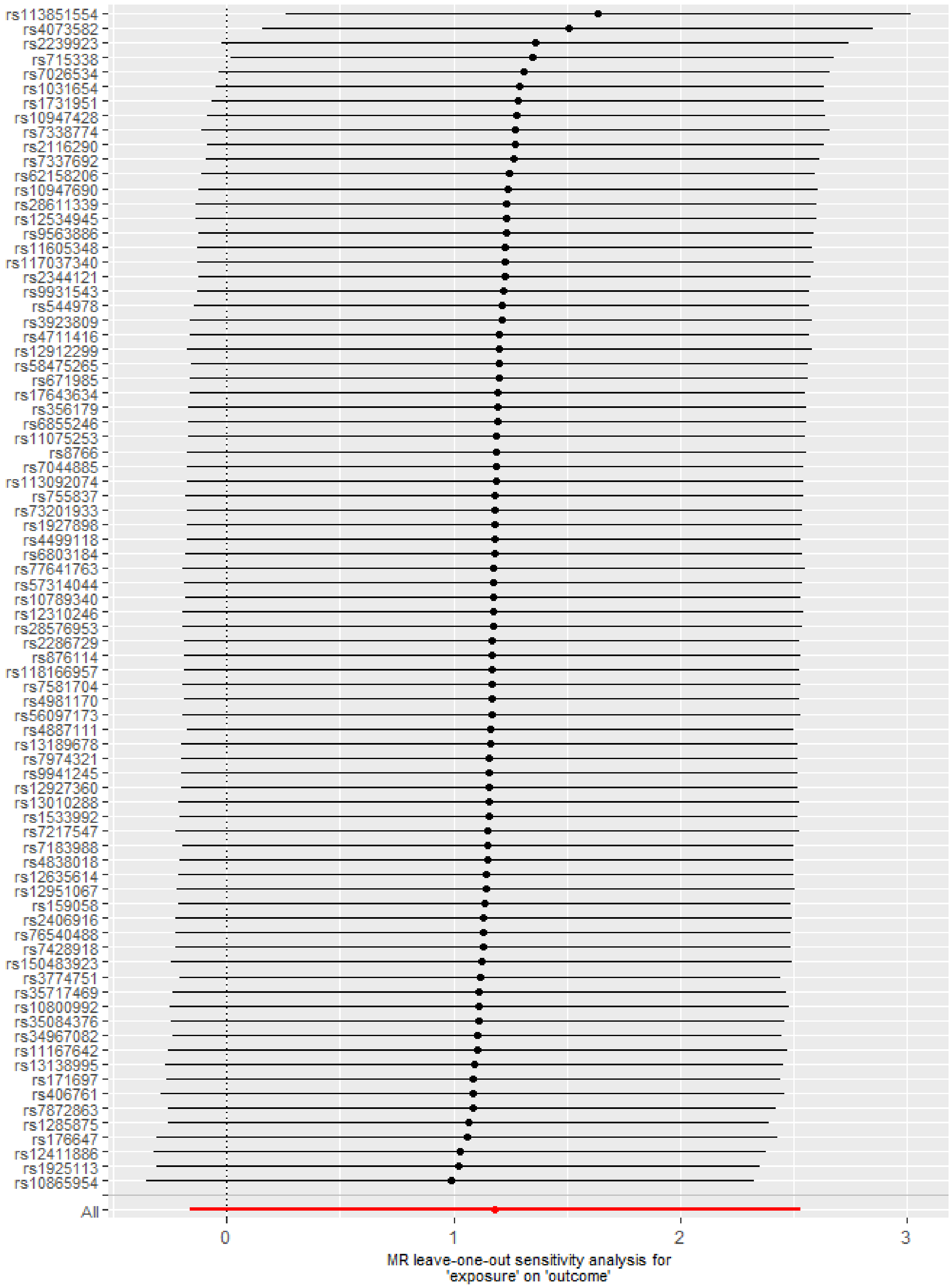

(f) Preterm birth

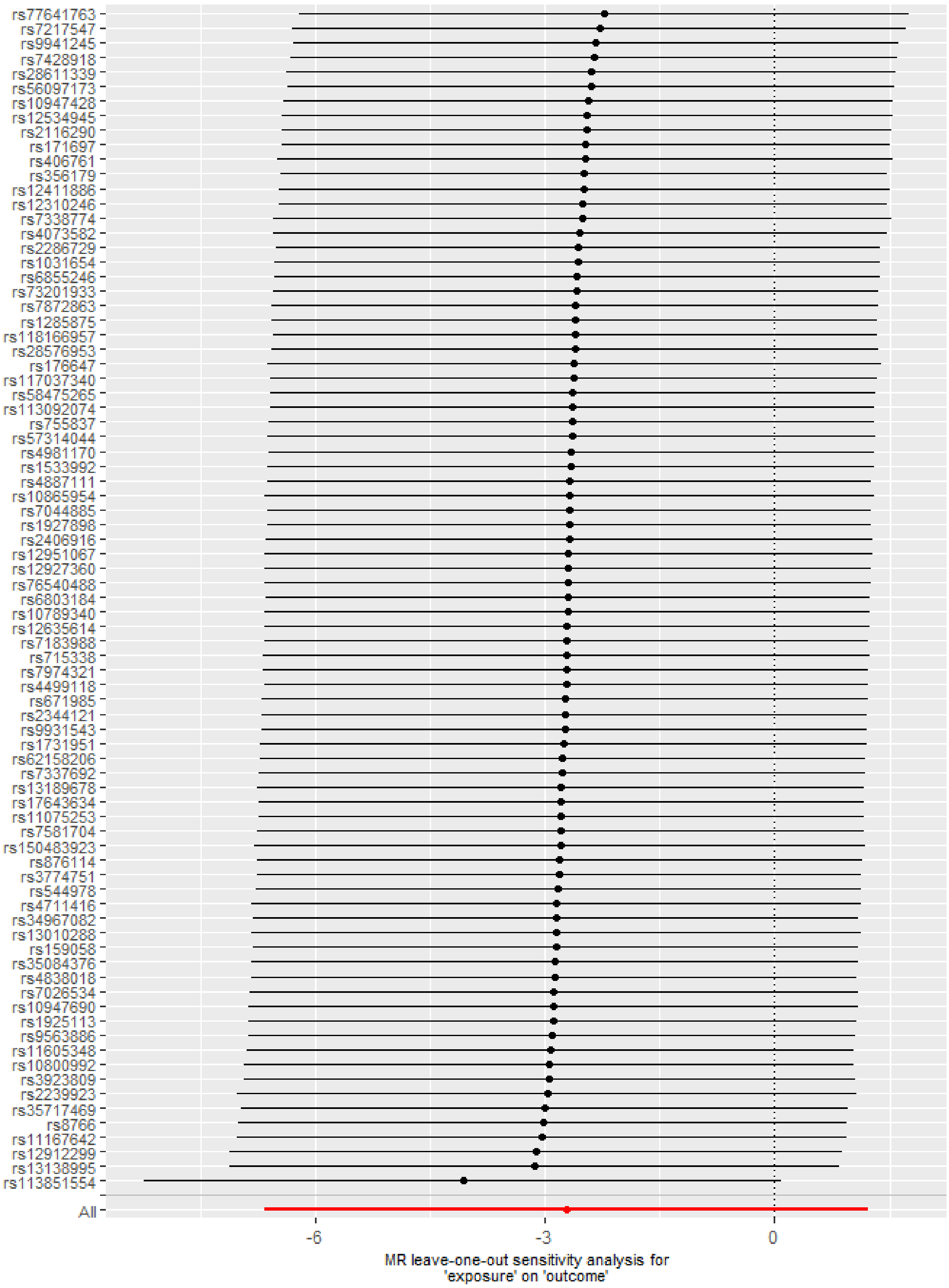

(g) Low offspring birthweight

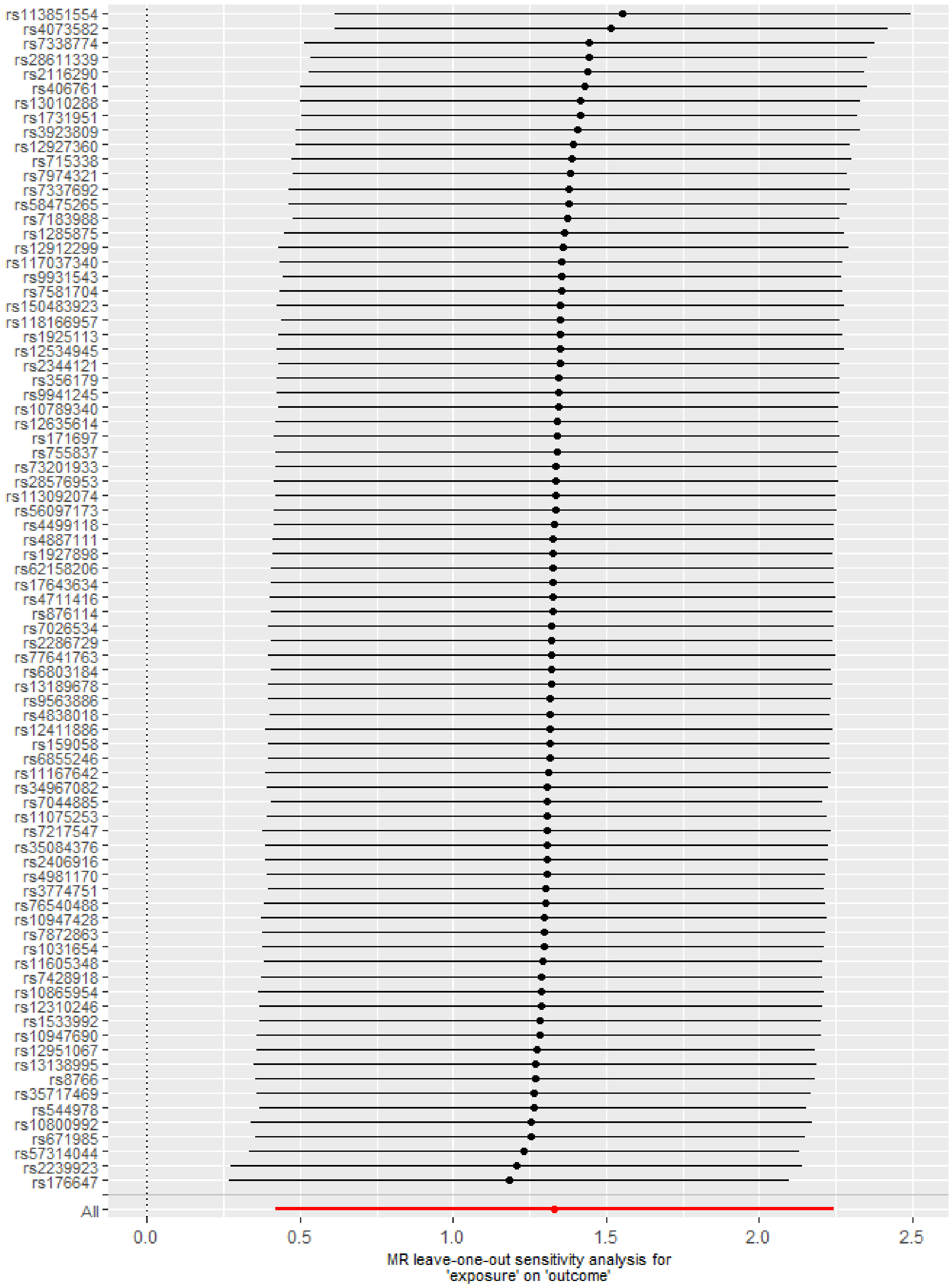

(h) High offspring birthweight

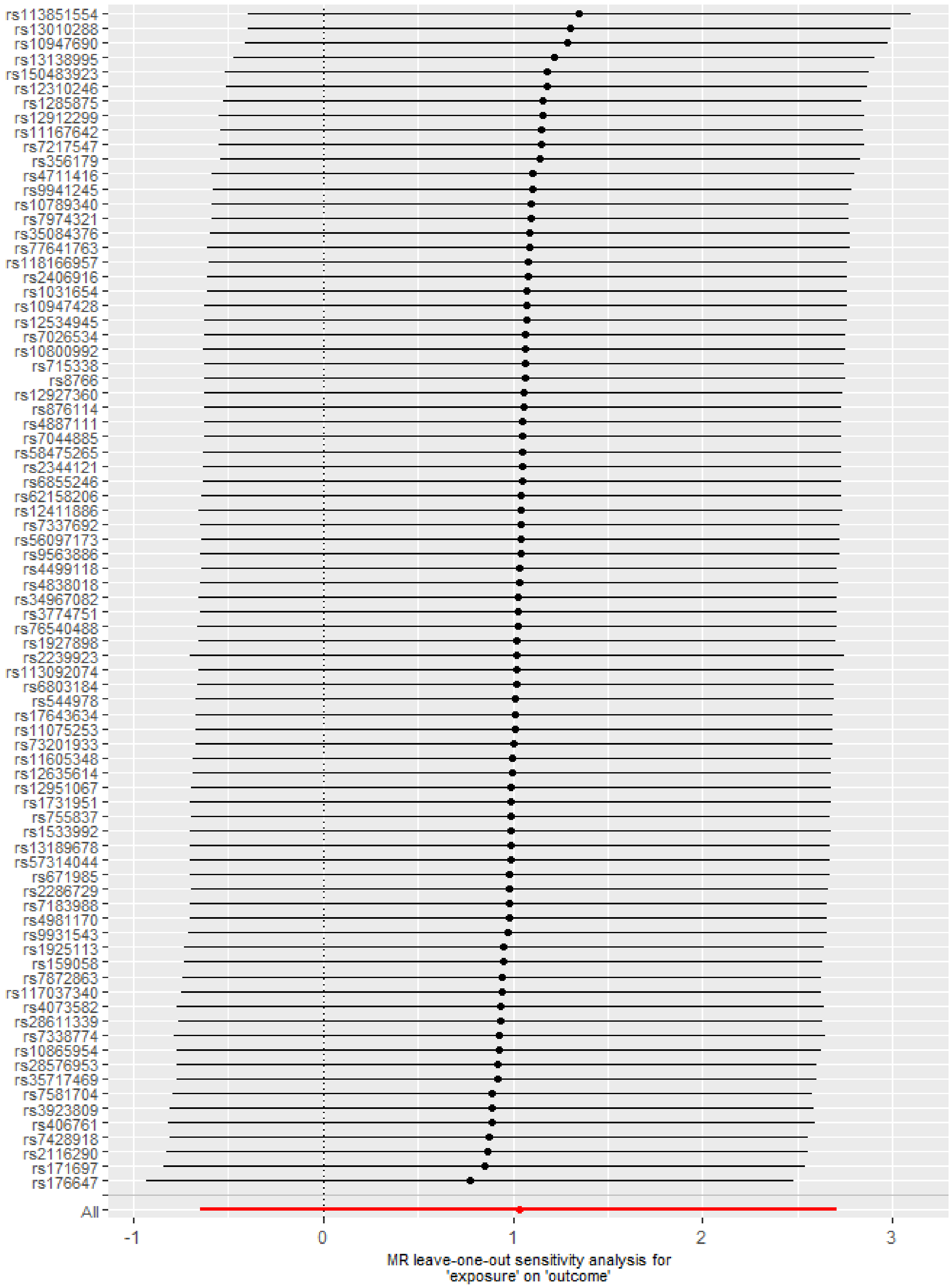

**S4 Fig. Leave-one (single nucleotide polymorphisms)-out sensitivity analysis for insomnia on pregnancy and perinatal outcomes in UK Biobank (dataset B on dataset A)**

(a) Stillbirth

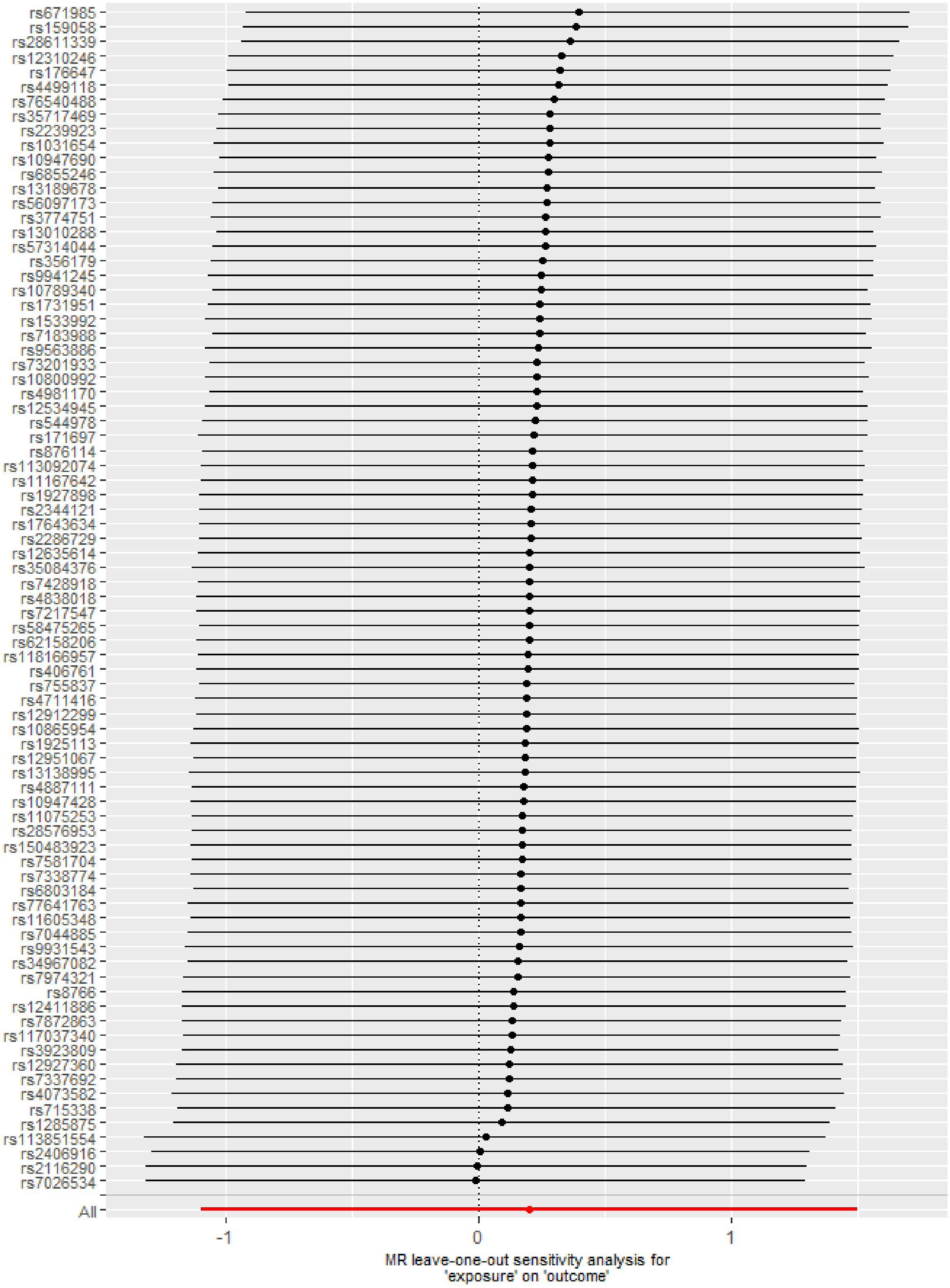

(b) Miscarriage

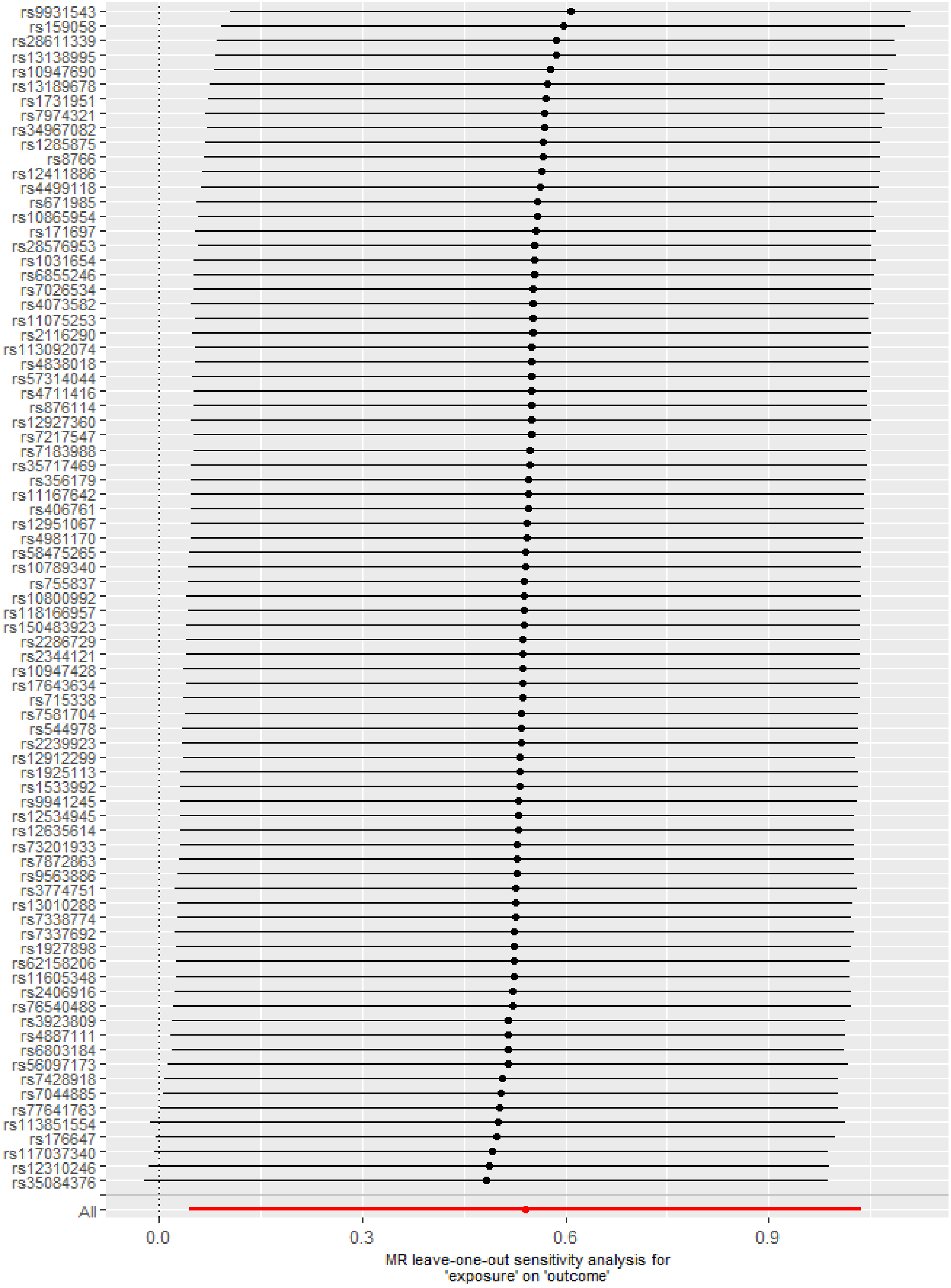

(c) Gestational diabetes

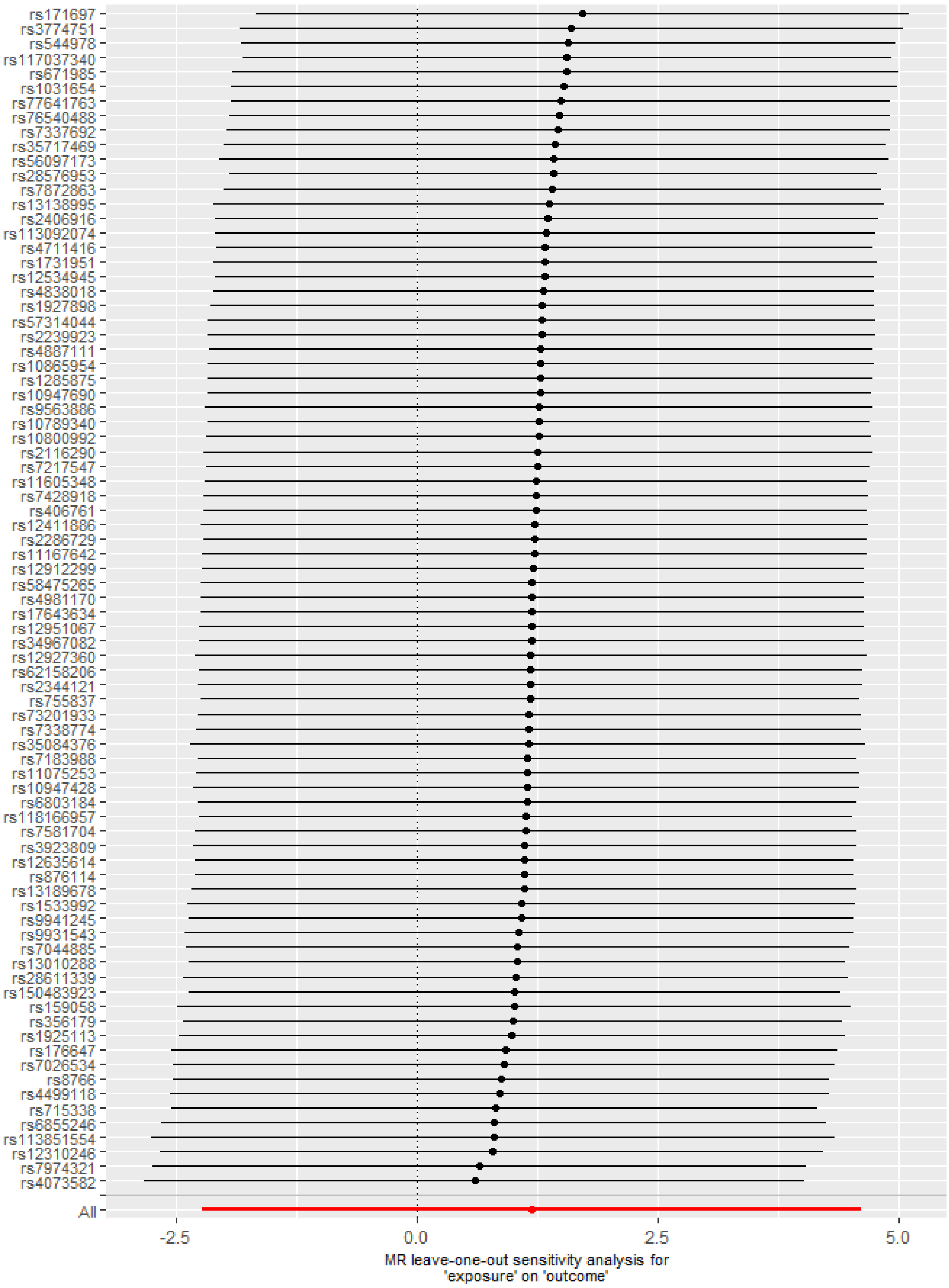

(d) Hypertensive disorders of pregnancy

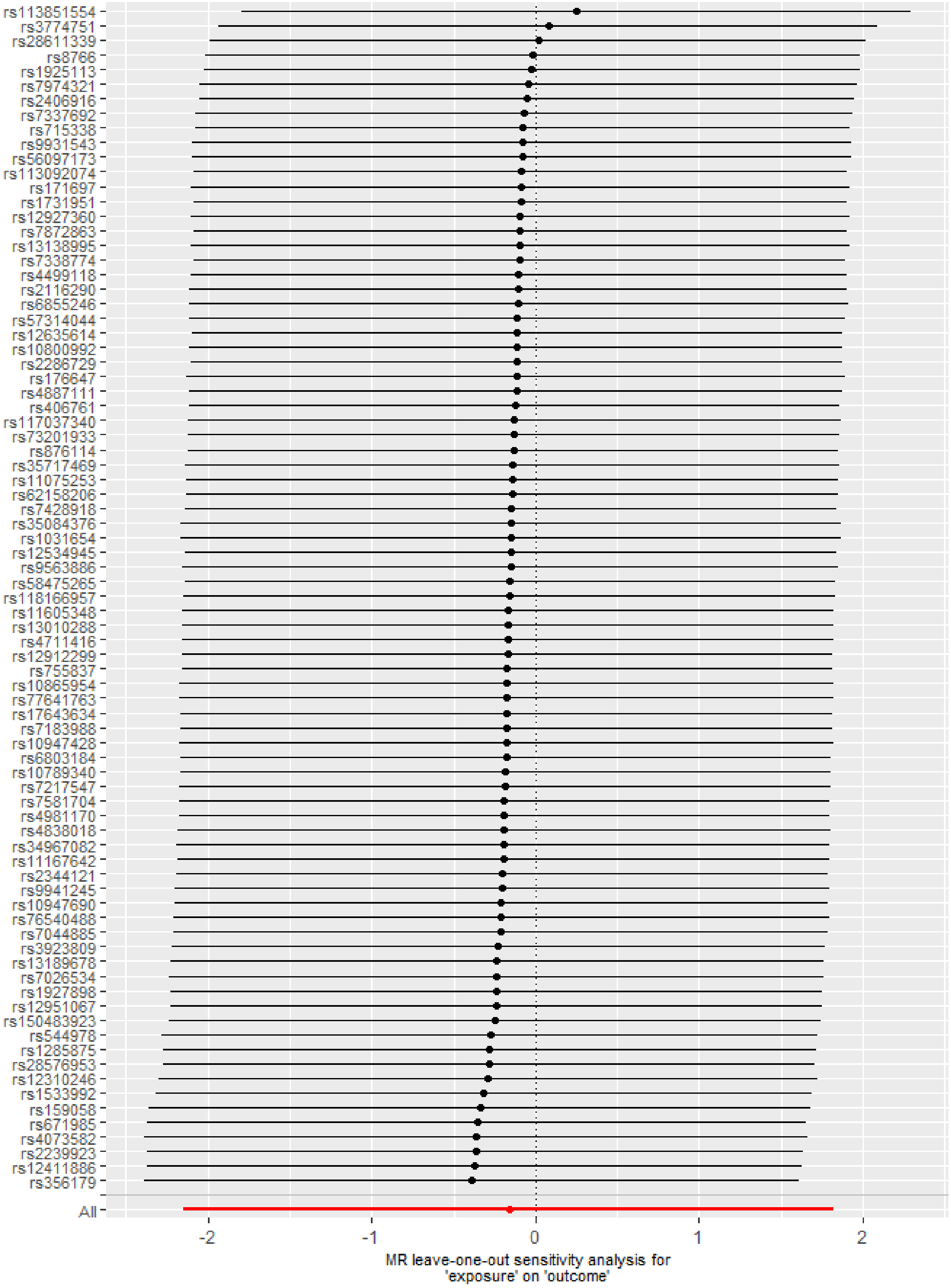

(e) Perinatal depression

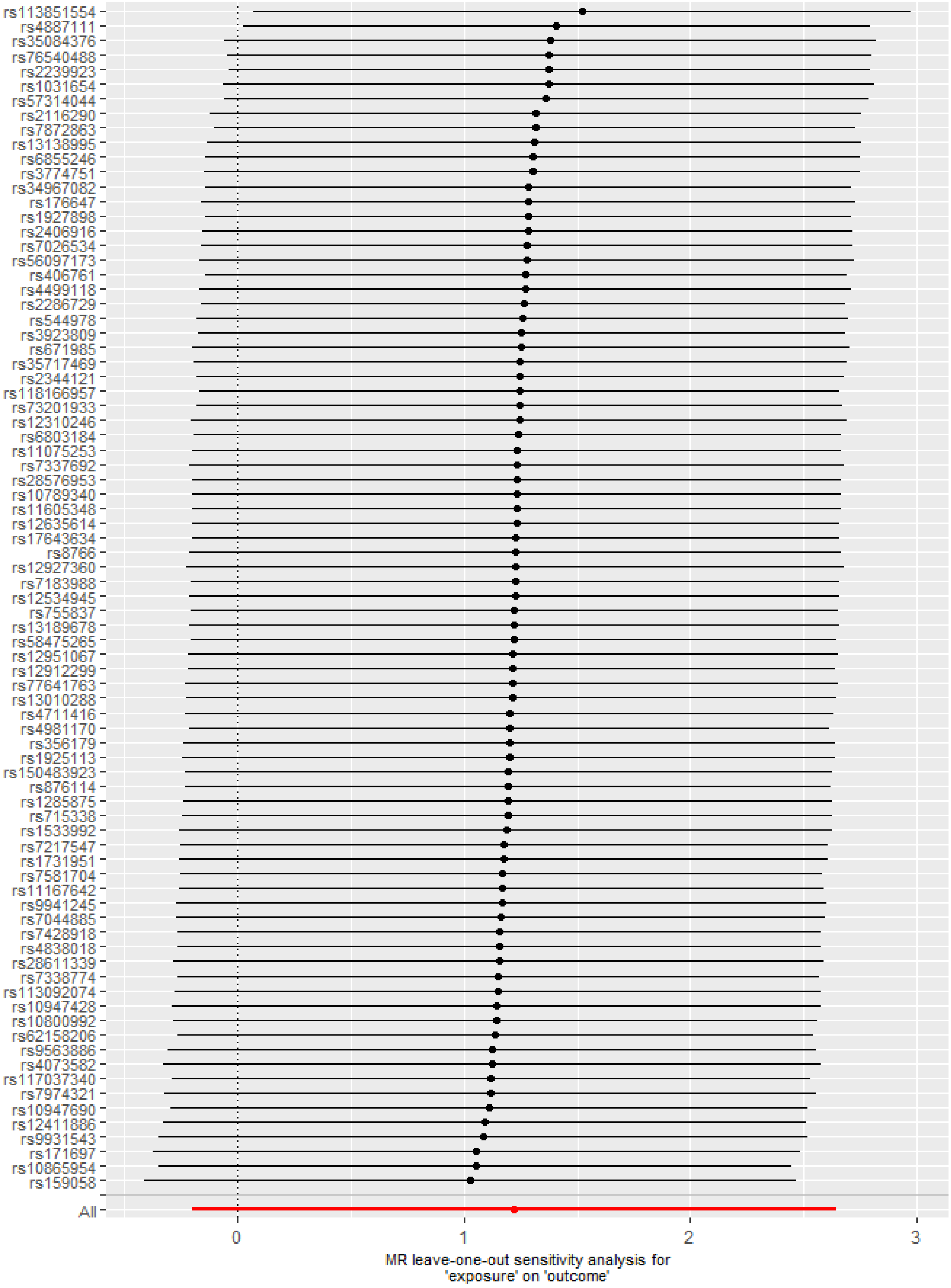

(f) Preterm birth

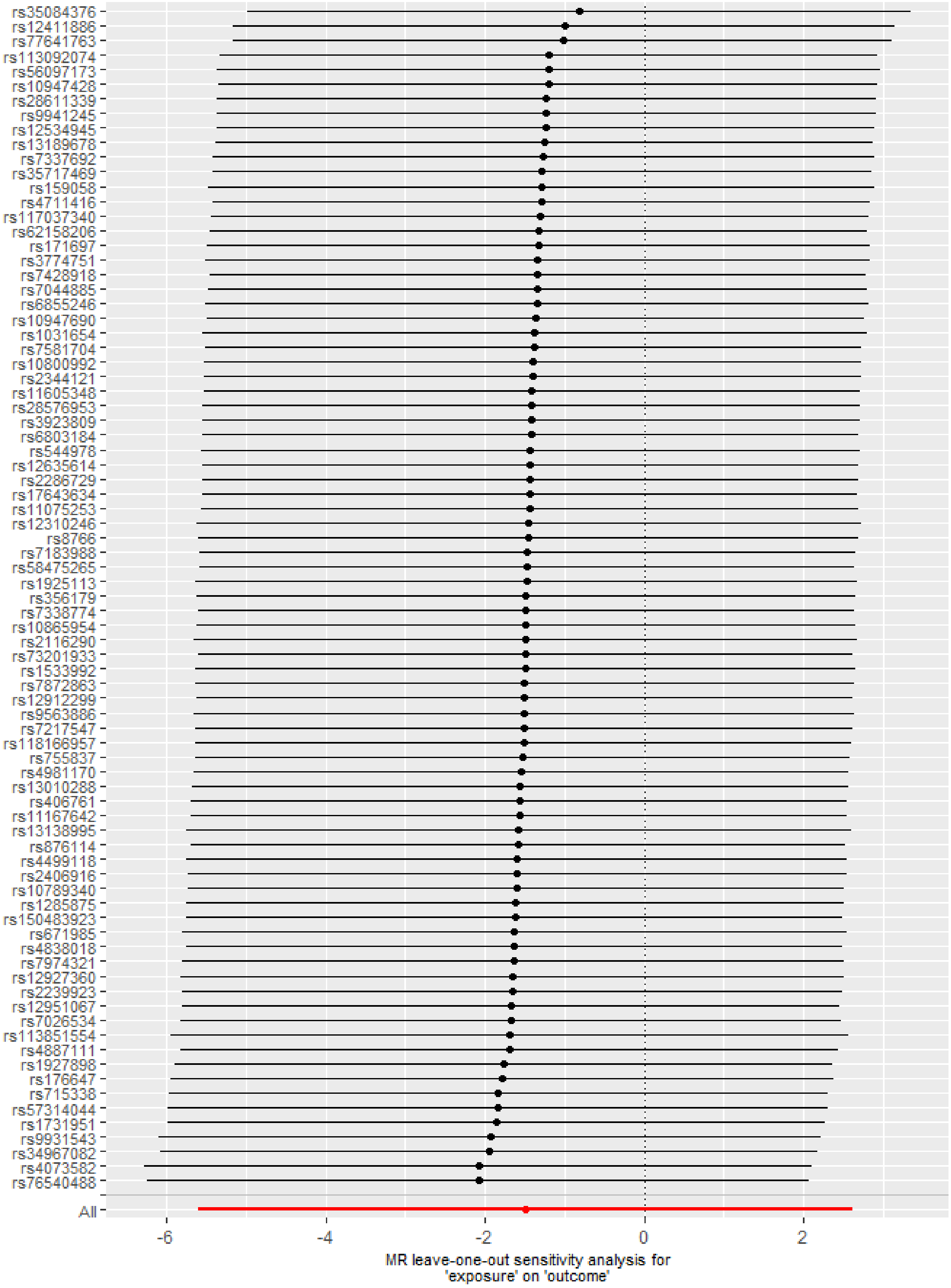

(g) Low offspring birthweight

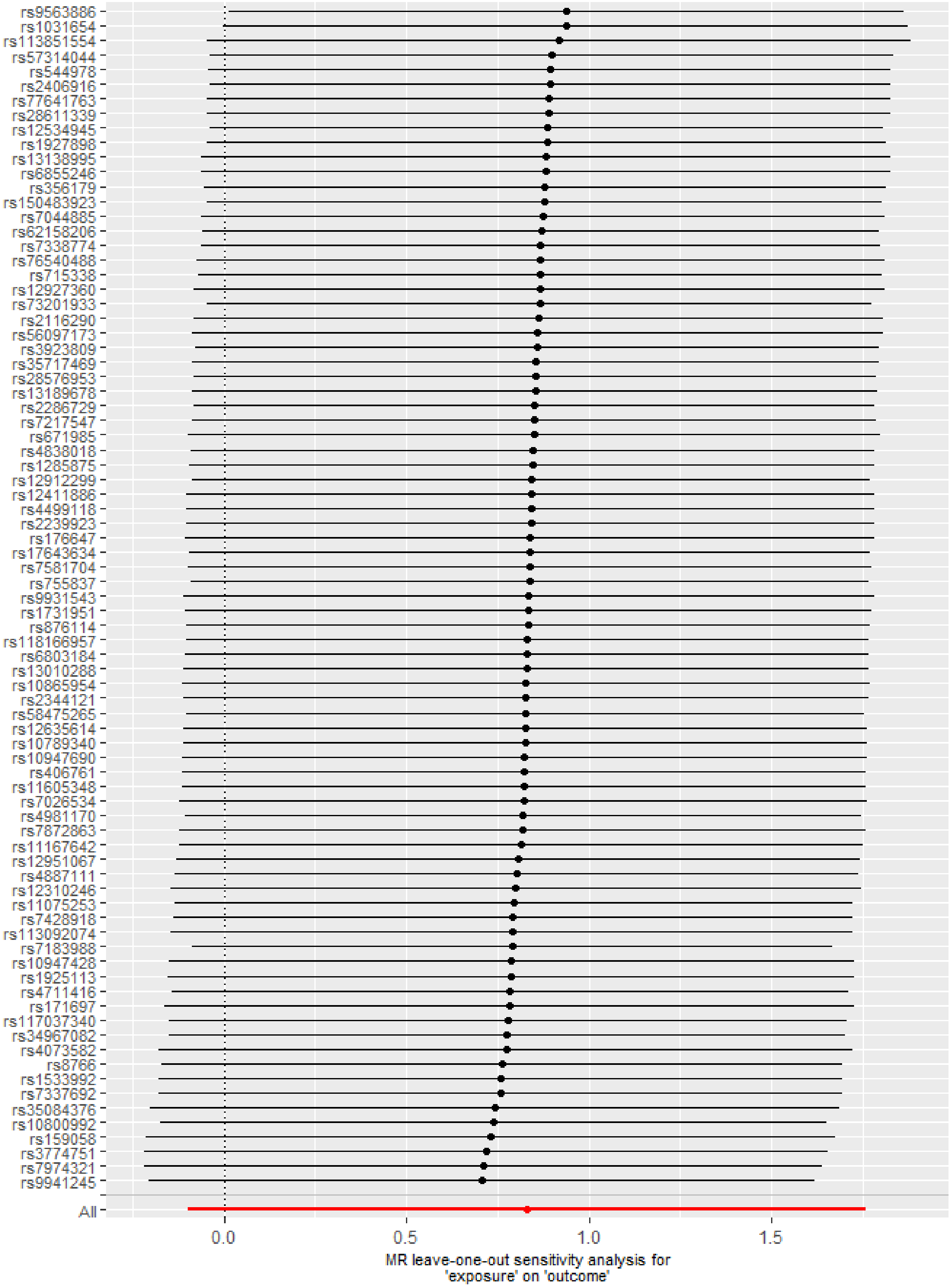

(h) High offspring birthweight

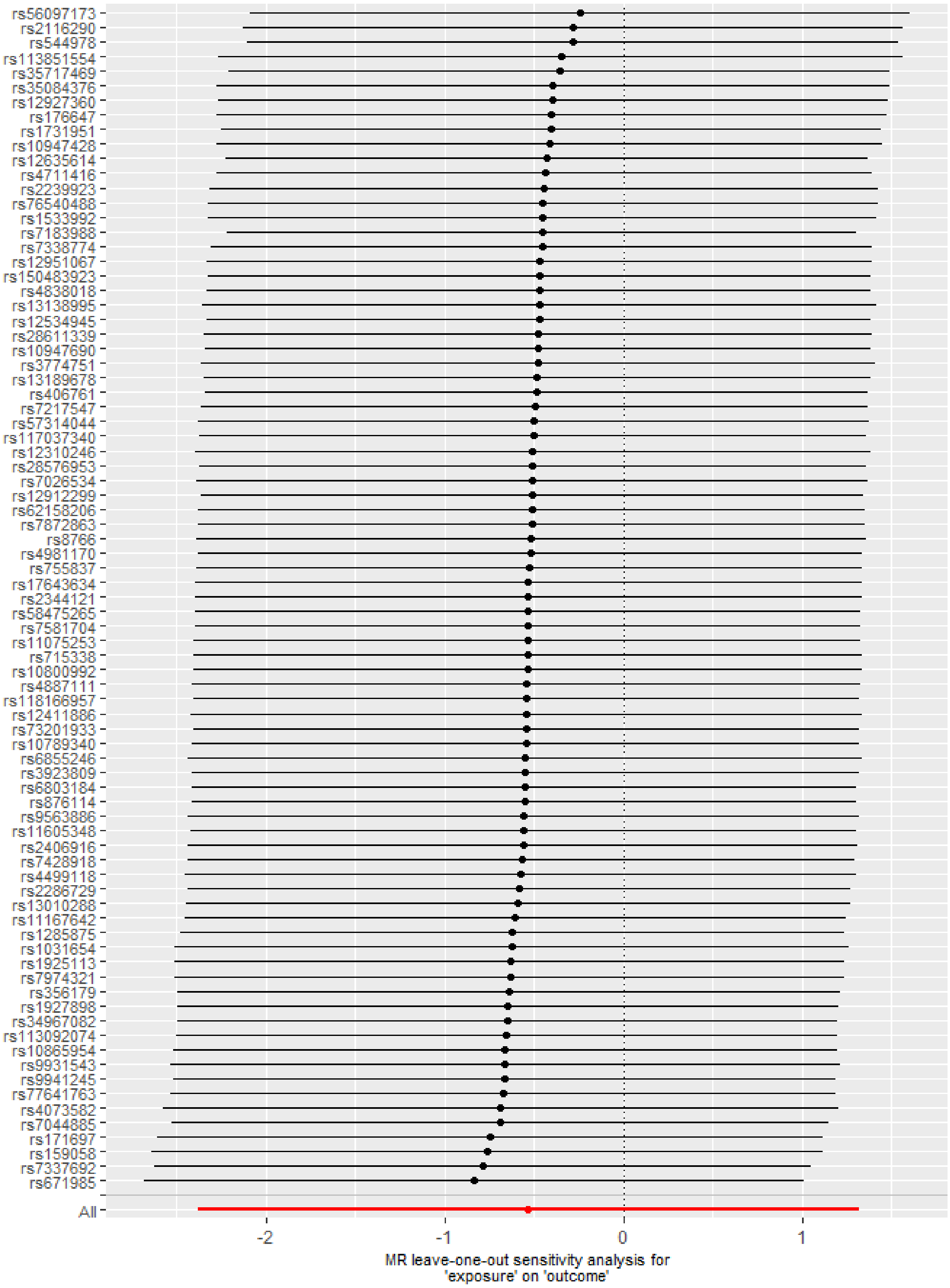

**S5 Fig. Leave-one (single nucleotide polymorphisms)-out sensitivity analysis for insomnia on pregnancy and perinatal outcomes in Avon Longitudinal Study of Parents and Children (ALSPAC), Born in Bradford (BiB), The Norwegian Mother, Father and Child Cohort Study (MoBa), and FinnGen**

(a) Stillbirth

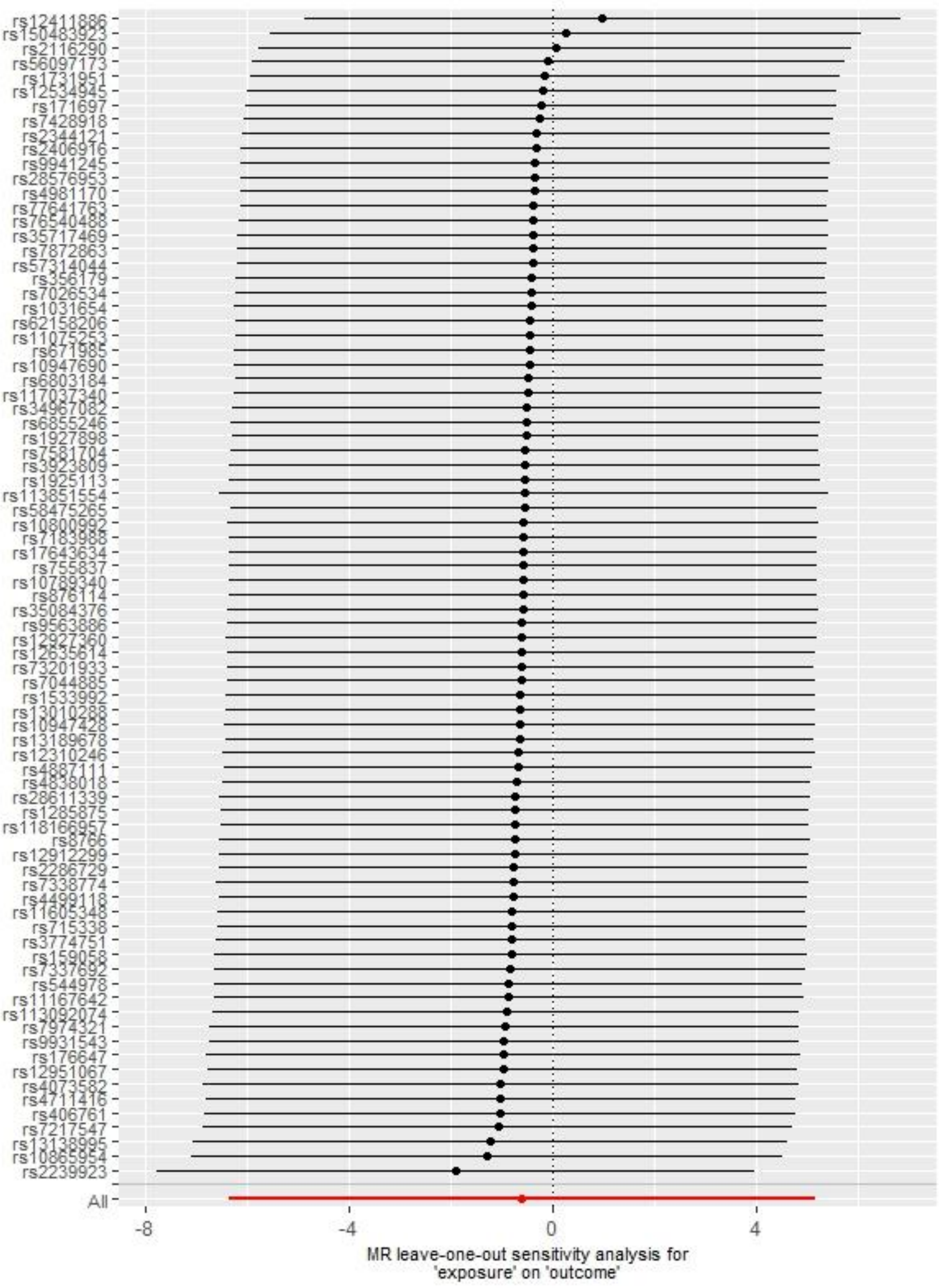

*This analysis only included ALSPAC, BiB and MoBa.

(b) Miscarriage

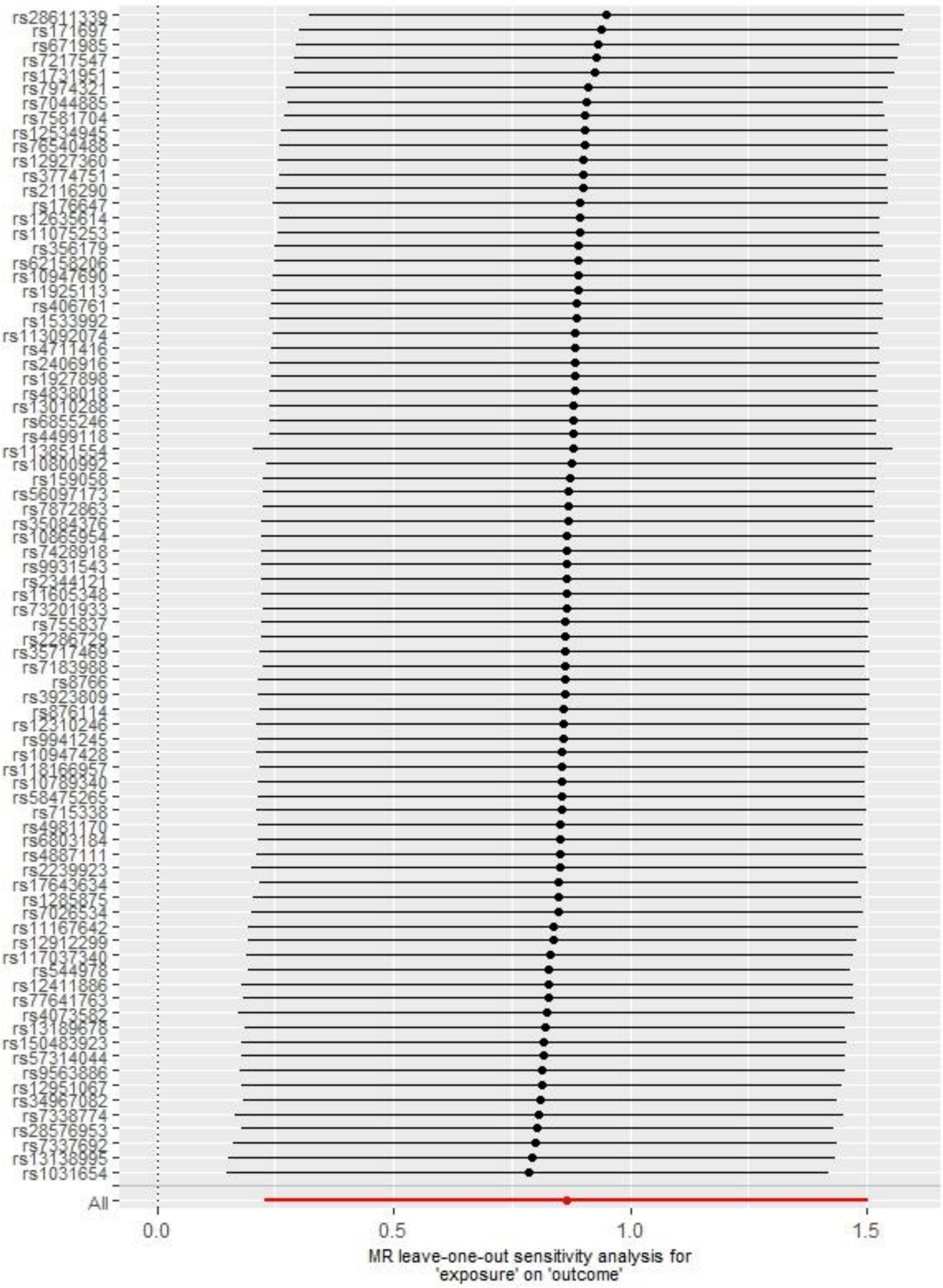


*This analysis included ALSPAC, BiB, MoBa and FinnGen.

(c) Gestational diabetes

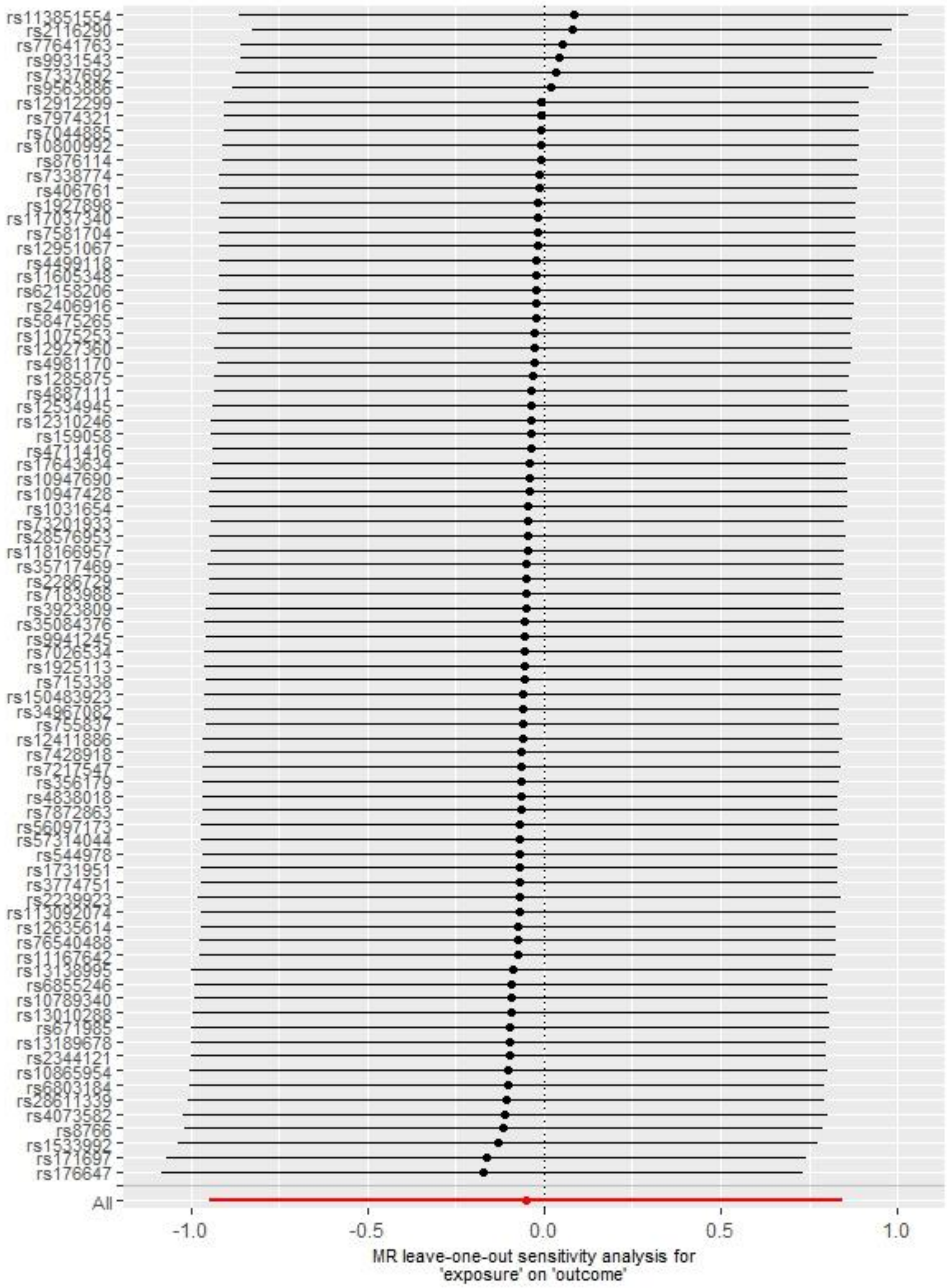


*This analysis included ALSPAC, BiB, MoBa and FinnGen.

(d) Hypertensive disorders of pregnancy

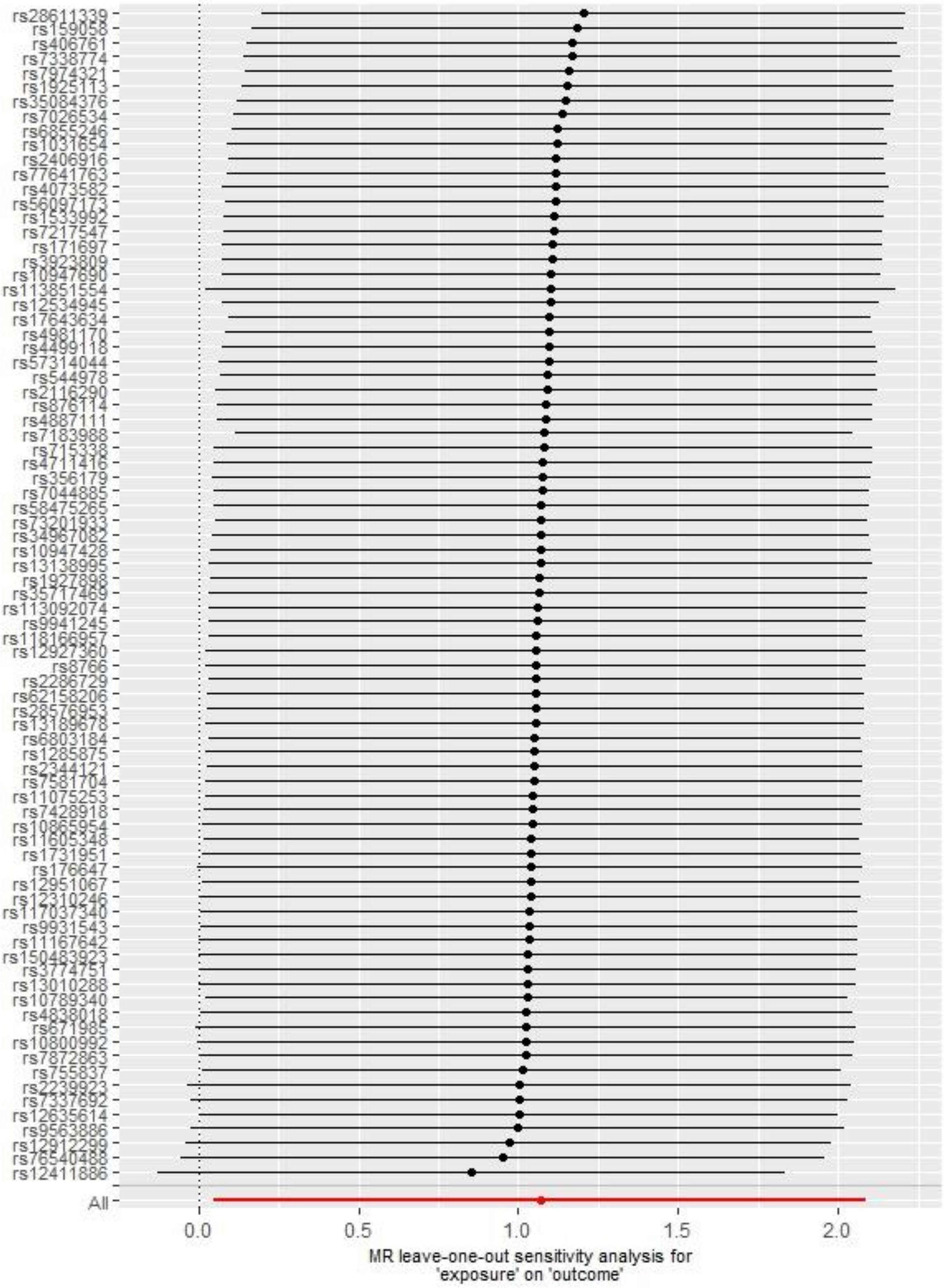


*This analysis included ALSPAC, BiB, MoBa and FinnGen.

(e) Perinatal depression

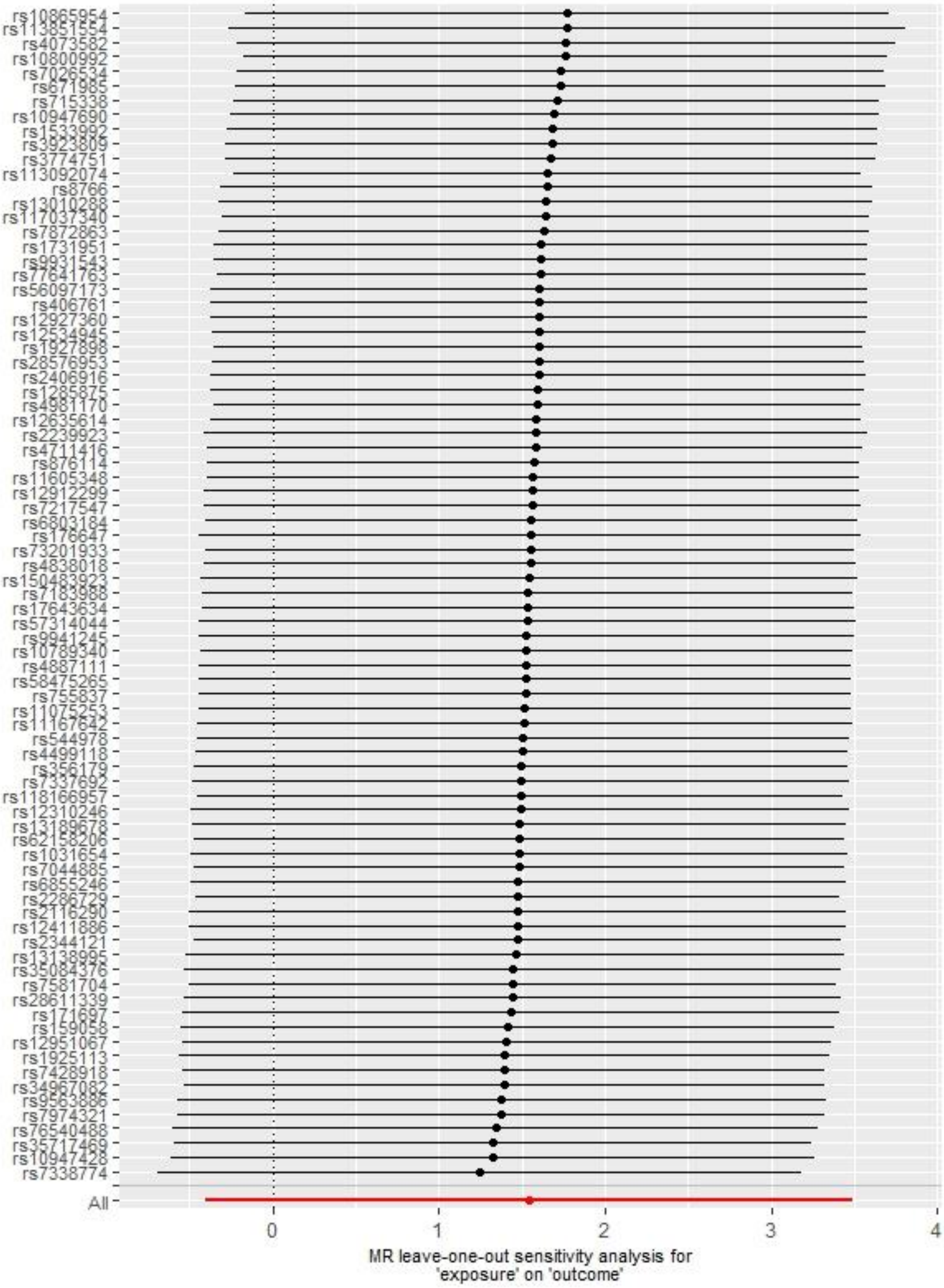


*This analysis only included ALSPAC, BiB and MoBa.

(f) Preterm birth

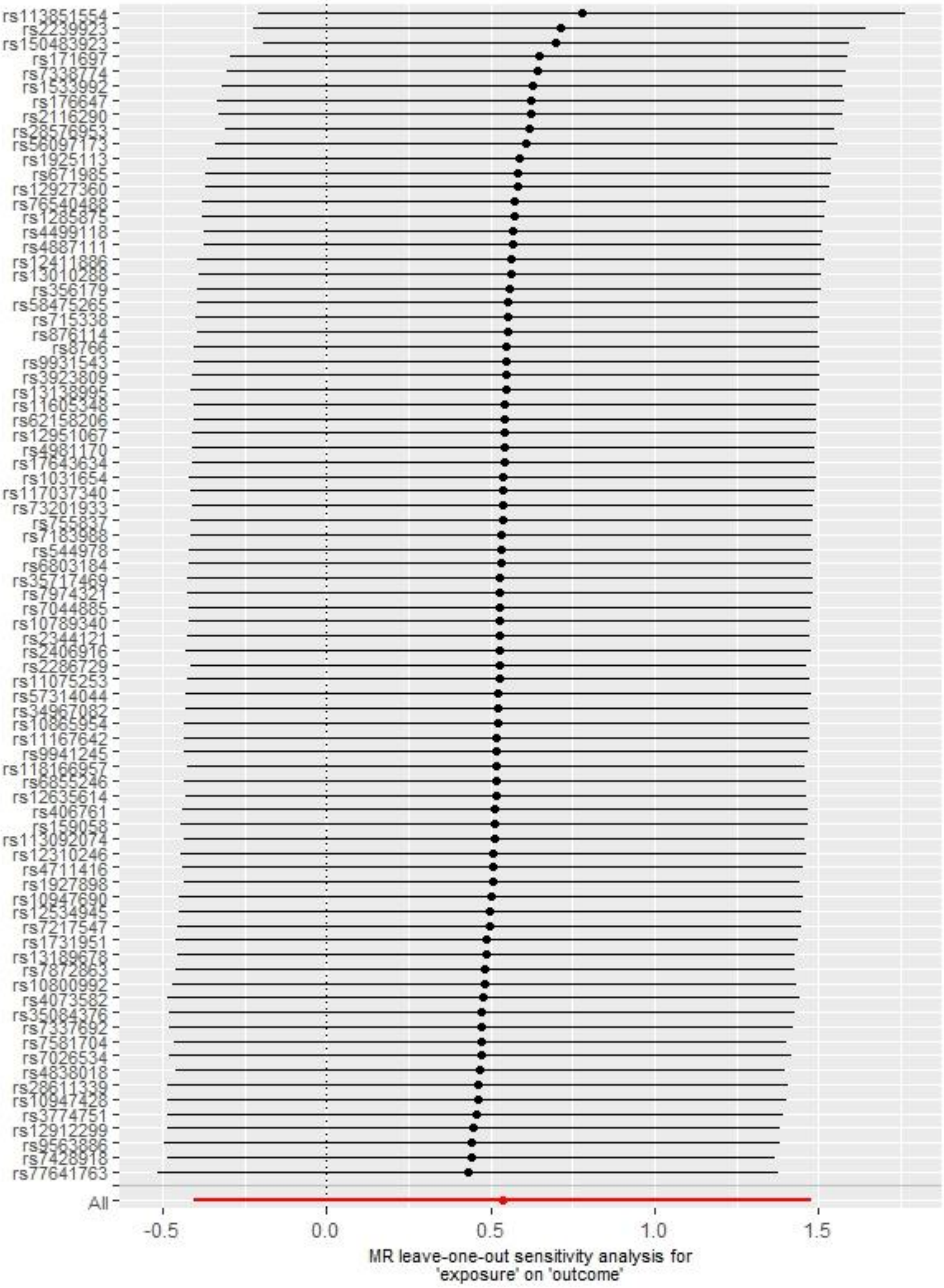


*This analysis included ALSPAC, BiB, MoBa and FinnGen.

(g) Low offspring birthweight

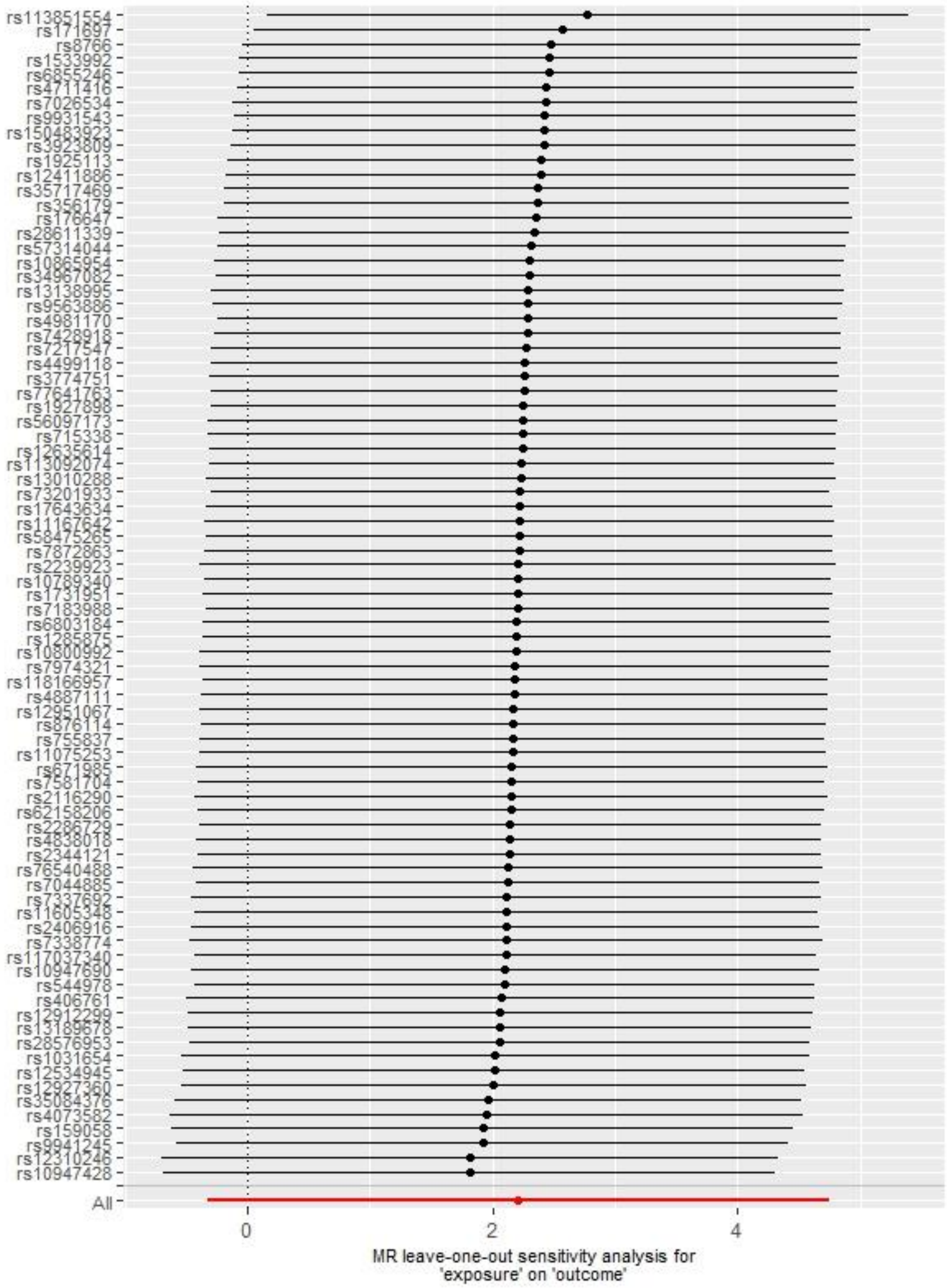


*This analysis only included ALSPAC, BiB and MoBa.

(h) High offspring birthweight

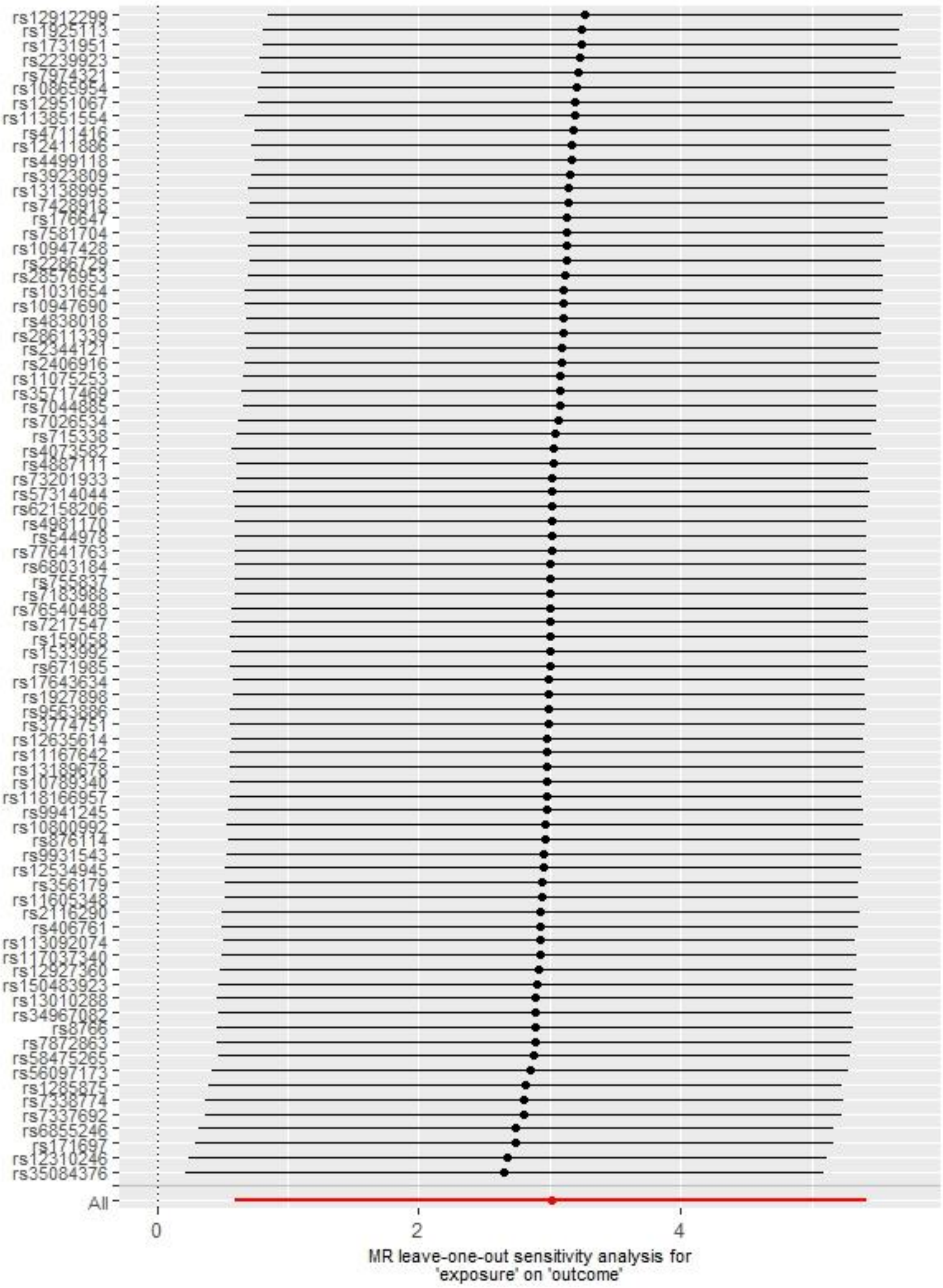


*This analysis only included ALSPAC, BiB and MoBa.

**S6 Fig. Associations of 81 maternal SNPs with pregnancy and perinatal outcomes comparing unadjusted to adjusted for fetal genotypes (N=18,663 mother-offspring pairs from three birth cohorts)**

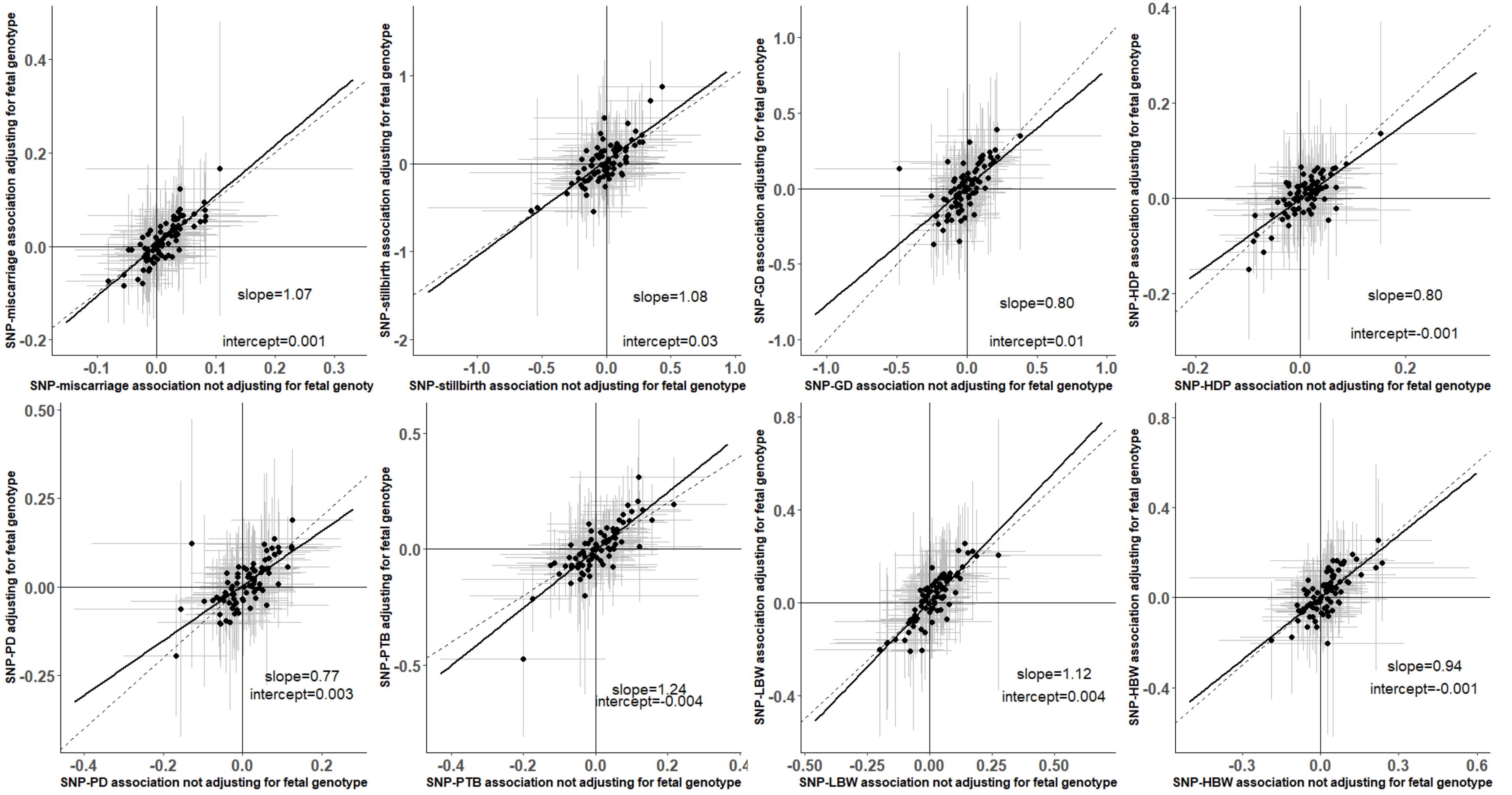


Black dots represent coordinates of ln (odds ratio) without fetal genotype adjustment (x-axis) and with that adjustment (y-axis) and the grey lines represent their 95% confidence intervals. Black solid lines are the fitted linear regression lines through the with and without fetal genotype adjustment ln (odds ratio). The black dash lines represent the line of perfect agreement between the ln (odds ratio) with and without fetal genotype adjustment.
Abbreviations: GD, gestational diabetes; HBW, high offspring birthweight; HDP, hypertensive disorders of pregnancy; LBW, low offspring birthweight; PD, perinatal depression; PTB, preterm birth; SNPs, single nucleotide polymorphisms.
